# Airway antibodies emerge according to COVID-19 severity and wane rapidly but reappear after SARS-CoV-2 vaccination

**DOI:** 10.1101/2020.11.25.20238592

**Authors:** Alberto Cagigi, Meng Yu, Björn Österberg, Julia Svensson, Sara Falck-Jones, Sindhu Vangeti, Eric Åhlberg, Lida Azizmohammadi, Anna Warnqvist, Ryan Falck-Jones, Pia C Gubisch, Mert Ödemis, Farangies Ghafoor, Mona Eisele, Klara Lenart, Max Bell, Niclas Johansson, Jan Albert, Jörgen Sälde, Deleah Pettie, Michael Murphy, Lauren Carter, Neil P King, Sebastian Ols, Johan Normark, Clas Ahlm, Mattias Forsell, Anna Färnert, Karin Loré, Anna Smed-Sörensen

**Author notes:** **Correspondence to:** Karin Loré and Anna Smed-Sörensen, Division of Immunology and Allergy, Department of Medicine Solna, Karolinska Institutet, Visionsgatan 4, BioClinicum J7:30, Karolinska University Hospital, 171 64 Stockholm, Sweden., E-mail addresses. Equal contribution (Karin Loré, Anna Smed-Sörensen).

## Abstract

Understanding the presence and durability of antibodies against SARS-CoV-2 in the airways is required to provide insights on the ability of individuals to neutralize the virus locally and prevent viral spread. Here, we longitudinally assessed both systemic and airway immune responses upon SARS-CoV-2 infection in a clinically well-characterized cohort of 147 infected individuals representing the full spectrum of COVID-19 severity; from asymptomatic infection to fatal disease. In addition, we evaluated how SARS-CoV-2 vaccination influenced the antibody responses in a subset of these individuals during convalescence as compared to naïve individuals. Not only systemic but also airway antibody responses correlated with the degree of COVID-19 disease severity. However, while systemic IgG levels were durable for up to 8 months, airway IgG and IgA had declined significantly within 3 months. After vaccination, there was an increase in both systemic and airway antibodies, in particular IgG, often exceeding the levels found during acute disease. In contrast, naïve individuals showed low airway antibodies after vaccination. In the former COVID-19 patients, airway antibody levels were significantly elevated after the boost vaccination, highlighting the importance of prime and boost vaccination also for previously infected individuals to obtain optimal mucosal protection.

## Introduction

Severe acute respiratory syndrome coronavirus 2 (SARS-CoV-2) infection that causes coronavirus disease 2019 (COVID-19) presents with a wide range of disease severity from asymptomatic to fatal (1, 2). Individuals of advanced age and/or those with comorbidities are overrepresented among patients who develop severe disease (3). However, the majority of SARS-CoV-2 infected individuals experience asymptomatic infection or only mild disease (4).

Systemic antibodies against the SARS-CoV-2 nucleocapsid (N) and the viral surface glycoprotein spike (S) as well as against the receptor binding domain (RBD) (5, 6) of the S protein have been studied extensively (7-11). Responses against the internal N protein are often readily detectable but their contribution to protection and control of disease is not clear (8, 10). In contrast, antibody responses against S and, in particular, against the RBD result in virus neutralization (12). Responses against the RBD are thus likely necessary for protection from re-infection or prevention of symptomatic disease. However, the presence and durability of antibodies during COVID-19 in the airways is still not well understood.

The respiratory tract is the initial site of viral infection and replication. The availability of antibodies at this site could therefore determine the ability to neutralize the virus locally in case of (re-) exposure and prevent viral spread. Generally, antibodies present in the circulation and at local sites are the result of secretion from short-lived plasmablasts and/or terminally differentiated plasma cells in the bone marrow or mucosal sites (13). However, the response to a secondary infection once antibody titers have waned below protective levels mostly relies on the presence of resting antigen-specific memory B cells that are rapidly activated upon antigen re-exposure (13). Whether vaccination against SARS-CoV-2 also elicits systemic antibody responses in addition to local antibodies in the airways of individuals who recovered from COVID-19, and via which mechanism, is currently unknown.

In this study we present data from a cohort of patients that we have followed since mid-March 2020, which was the start of the pandemic in Sweden. We show longitudinal data on virus-specific systemic and airway antibody and B cell memory responses generated in this clinically well-characterized cohort of individuals with SARS-CoV-2 infection (n=147) ranging from asymptomatic SARS-CoV-2 infection to fatal COVID-19 disease. In addition, we show how subsequent SARS-CoV-2 vaccination during the convalescent phase significantly boosts not only the systemic but also airway antibody responses.

## Results

### Patient enrollment, assessment of disease severity and timeline

Individuals were sampled longitudinally in blood and airways during acute infection/symptomatic disease and during convalescence (median 3 and 8 months from symptom onset). Donor-matched plasma, peripheral blood mononuclear cells (PBMC), nostril swabs (NSW) and nasopharyngeal aspirates (NPA) were collected from all patients across disease severities whereas endotracheal aspirates (ETA) were only collected from intubated patients receiving intensive care (Figure 1). Disease severity was assessed daily, using a seven-point scale derived from the respiratory domain of the sequential organ failure assessment (SOFA) score (14, 15), with additional levels for non-admitted and fatal cases (Table 1). Patients were grouped based on peak disease severity, which may differ from disease severity at time of sampling (Table 1 and Figure 1B). In addition, pre-pandemic healthy controls (PPHC) (n=30) as well as individuals with influenza-like symptoms, and possible SARS-CoV-2 exposure, but with negative diagnostic PCR results (PCR-) (n=9) were sampled in the same way and included as controls. Generally, severe patients were sampled later after symptom onset as compared with individuals with mild disease resulting in a large time frame of study inclusion with respect to symptom onset (Table 1 and Figure 1B) (16). For simplicity, the sampling period/study inclusion during ongoing infection and hospitalization (for those hospitalized) is referred to as the “acute” phase. Samples collected at the first follow-up visit during convalescence (range 46-168 days from symptom onset; median 108 days, coefficient of variation 21.56%) are referred to as the “3 months” timepoint whereas those collected at the second follow-up visit (range 187-344 days; median 245 days, coefficient of variation 9.52%) are referred to as the “8 months” timepoint. Time of the first convalescent follow-up sampling from acute sampling ranged 33-159 days; median 90 days, coefficient of variation 24.84% (Table 1).

**Figure 1.**
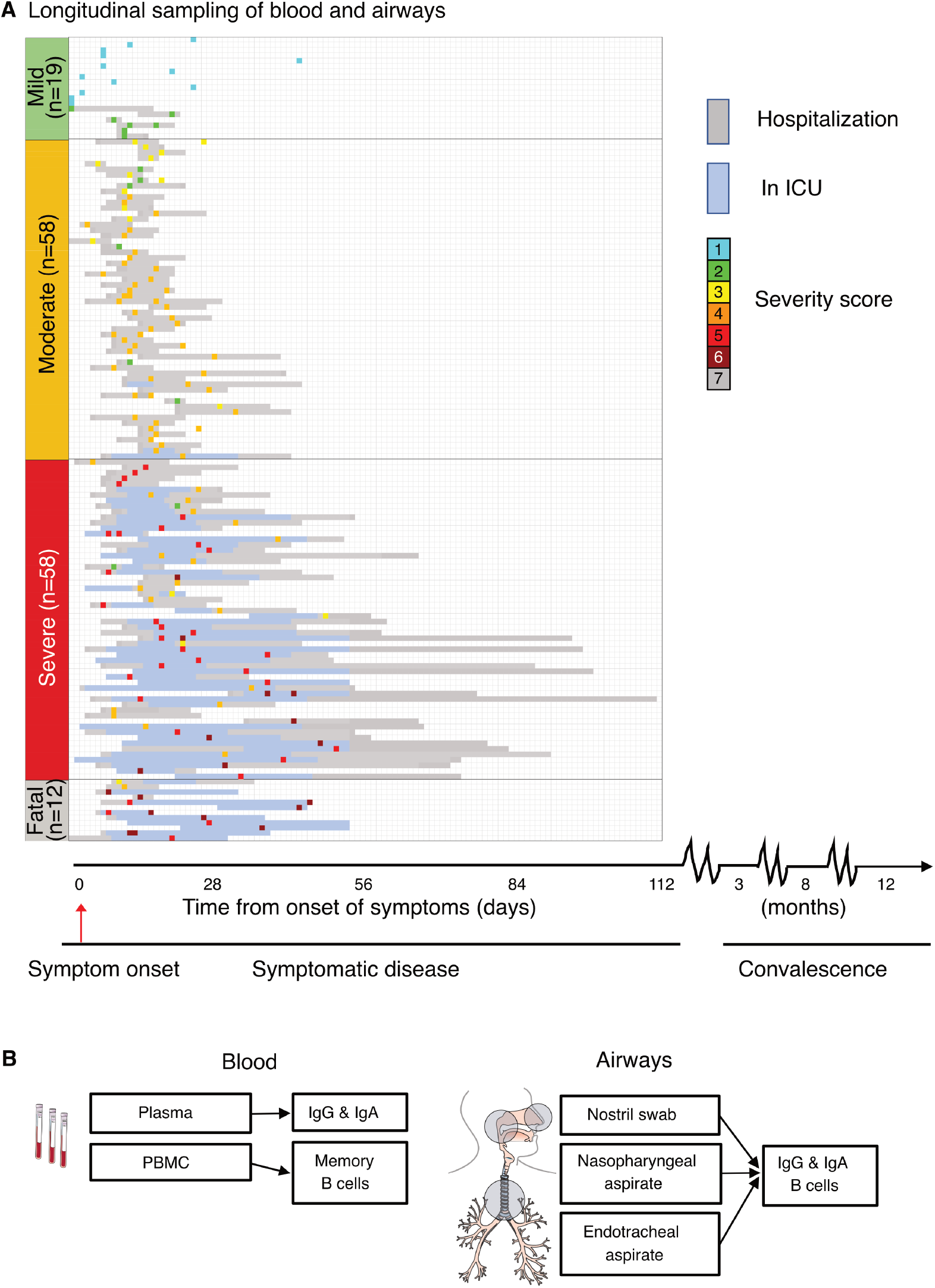
Study and sampling overview. **(A)** Overview of study cohort (n=147), timeline of longitudinal sampling, hospital admission/discharge, level of care and outcome for each patient. Patients are group based on peak disease severity (PDS); mild (PDS 1 and 2), moderate (PDS 3 and 4), severe (PDS 5 and 6) and fatal (PDS 7). Individual inclusion sample for each patient is color-coded based on disease severity at the time of sampling. **(B)** Overview of the anatomical compartments analyzed, and the measurements performed.

**Table 1.** Clinical characterization of the SARS-CoV-2 infected cohort

| Peak disease severity | 1 | 2 | 3 | 4 | 5* | 6* | 7 |
| --- | --- | --- | --- | --- | --- | --- | --- |
| Resp. SOFA score | 0 | 0 | 1 | 2 | 3 | 4 |  |
| Admitted (Y/N) | N | Y | Y | Y | Y | Y | Y |
| PFI (kPa) | > 53 | > 53 | < 53 | < 40 | < 27 | < 13 | - |
| SFI | > 400 | > 400 | ≤ 400 | ≤ 315 | ≤ 235 | < 150 |  |
| No. of individuals (%) | 13 (8.8) | 6 (4.1) | 10 (6.8) | 48 (33) | 19 (13) | 39 (27) | 12 (8.2) |
| Age, mean (Range) | 44 (24-72) | 60 (41-72) | 56 (46-78) | 55 (24-76) | 57 (42-74) | 61 (25-77) | 66 (52-78) |
| Male (%) | 5 (38) | 2 (33) | 6 (60) | 38 (79) | 15 (79) | 34 (87) | 9 (75) |
| Days from symptoms to admission – median (Range) | - | 10 (0-14) | 8.5 (4-14) | 10 (3-21) | 7 (2-14) | 10 (2-35) | 7 (1-28) |
| Days from symptoms to inclusion “Acute” – median (Range) | 9 (3-44) | 11 (0-20) | 13.5 (6-18) | 13 (4-32) | 21 (5-40) | 22 (7-54) | 13 (8-44) |
| Days from symptoms to 3-Mo follow-up – median (Range) | 102 (88-136) | 99,5 (82-103) | 112 (81-127) | 109 (46-155) | 109 (48-130) | 120 (53-168) | - |
| Days from symptoms to 8-Mo follow-up – median (Range) | 232 (187-264) | 238 (212-250) | 245 (227-303) | 247 (233-314) | 241 (220-270) | 254 (224-344) | - |
| VL (Ct value) median (Range) | 27.5 (40-14) | 25.0 (29-14) | 26.7 (36-15) | 26.8 (36-12) | 25.8 (36-19) | 24.0 (37-14) | 20.5 (32-13) |
| CCI, mean (SD) | 1 (2) | 2 (1) | 1 (1) | 2 (2) | 2 (1) | 2 (1) | 3 (1) |
| BMI, mean (SD) | 24.1 (4.5) | 25.1 (2.2) | 26.0 (3.2) | 30.3 (4.2) | 29.2 (5.3) | 28.6 (4.7) | 28.6 (2.4) |
| Hypertension (%) | 1 (8.3) | 0 (0) | 2 (20) | 20 (42) | 8 (42) | 15 (38) | 9 (75) |
| Diabetes (%) | 2 (17) | 0 (0) | 1 (10) | 14 (29) | 5 (26) | 9 (23) | 3 (25) |
| Current smokers (%) | 0 (0) | 0 (0) | 2 (20) | 5 (11) | 1 (5.3) | 2 (5.3) | 0 (0) |
| ACE-I (%) | 0 (0) | 0 (0) | 0 (0) | 5 (10) | 1 (5.3) | 4 (10) | 1 (9.1) |
| IS drugs (%) | 1 (7.7) | 0 (0) | 0 (0) | 4 (8.3) | 2 (11) | 5 (13) | 1 (8.3) |

Peak disease severity: 1-2 (Mild), 3-4 (Moderate), 5-6 (Severe), 7 (Fatal)
Resp. SOFA: Respiratory Sequential Organ Failure Assessment
PFI: PaO<sub>2</sub>/FiO<sub>2</sub>-index
SFI: SpO<sub>2</sub>/FiO<sub>2</sub>-index
VL: Viral Load
CCI: Charlson comorbidity index
BMI: body mass index
ACE-I: angiotensin converting enzyme-inhibitors
IS: immunosuppressive

### Plasma IgG and IgA responses to N, S and RBD across COVID-19 severity during acute disease and after recovery

We first assessed systemic IgG and IgA responses against N, S and RBD at the time of study inclusion that ranged between 0-54 days from onset of symptoms; median 16 days for the whole cohort (Table 1). Both IgG and IgA levels against all viral proteins followed the degree of disease severity with increasing levels in patients with mild, moderate and severe disease respectively (Figure 2A). In line with previous reports, IgG against N were the most elevated in patients who had severe disease or a fatal outcome (8, 10). The degree of disease severity also associated with the levels of systemic inflammation as indicated by the levels of C-reactive protein (CRP) in blood and by the neutrophil-lymphocyte ratio (NLR) (Figure 2B). Interestingly, the levels of neutrophils also specifically associated with disease severity (Figure 2D) and with all of the systemic antibody responses during acute disease (Figure 2D and Supplementary figure 1). The levels of IgG during acute disease, and to a lower extent IgA, against all tested antigens, exhibited a positive correlation with the days from onset of symptoms (Supplementary figure 2A). This difference in antibody titers over time might be slightly accentuated by the fact that in our cohort the patients with moderate/severe disease, and even fatal outcome, for whom we initially observed low IgG titers against RBD, had an early study inclusion (on average 13 days from onset of symptoms). In fact, these patients showed significantly higher titers later during the acute phase (on average 19 days) (Supplementary figure 2B-C). Nonetheless, patients with mild disease displayed lower levels of plasma IgG against RBD as compared with more severe patients, also when samples were taken after similar duration of symptoms (Supplementary figure 2D). After 3 months from symptom onset, the IgG levels remained high in the plasma of patients recovering from moderate and severe disease, while the levels had further increased in the individuals who had a mild disease (Figure 3A). However, despite this increase over time, the antibody levels in mild patients never reached the levels observed for moderate and severe patients or for those who had a fatal outcome (Figure 3 and Supplementary figure 3A).

**Figure 2.**
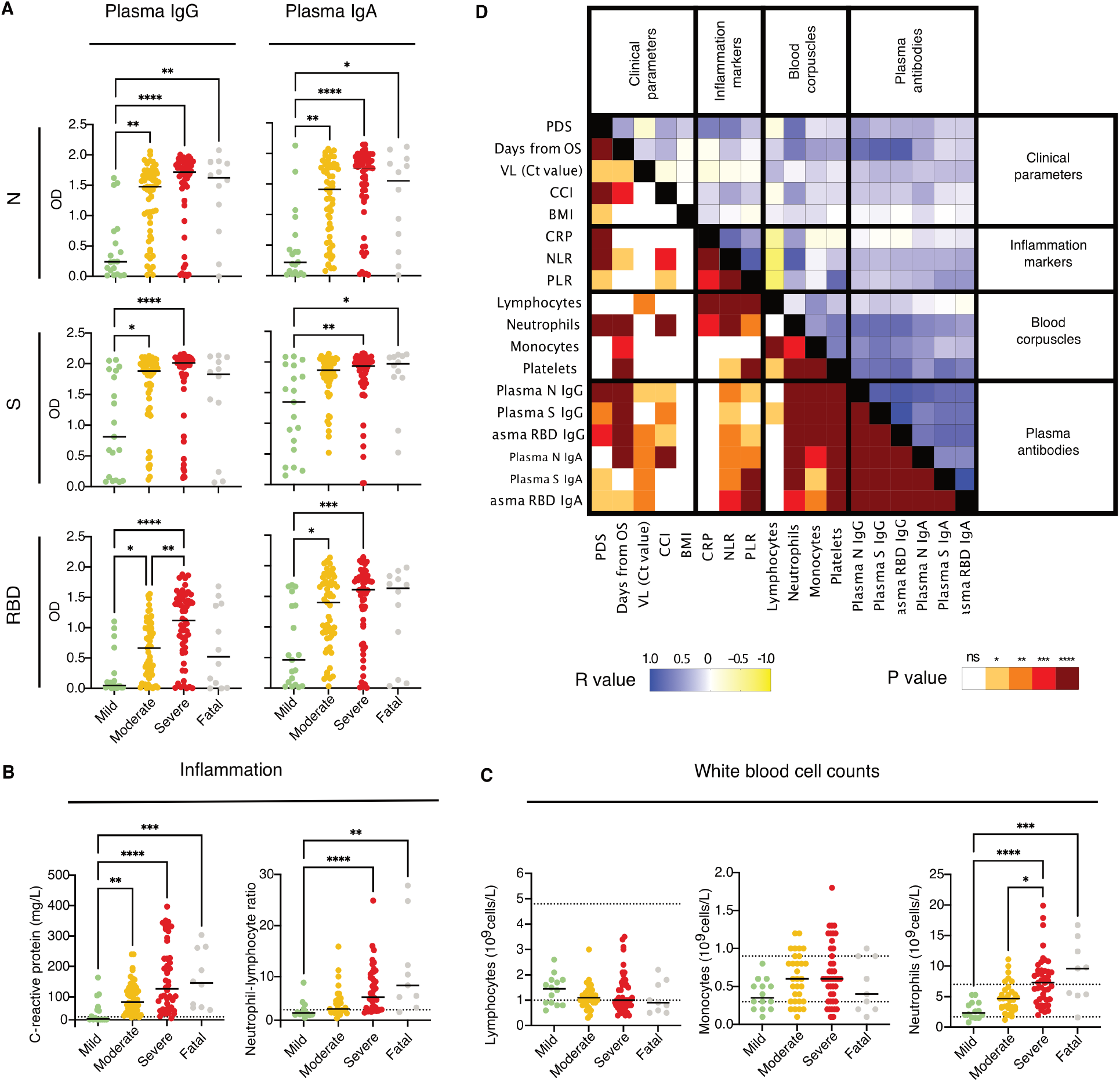
Systemic antibody responses, inflammation markers and other clinical parameters in relation to COVID-19 severity during acute disease. **(A)** Plasma IgG and IgA responses (n=19 for mild, n=58 for moderate, n=58 for severe and n=12 for fatal) against N, S and RBD are shown together with the levels of **(B)** C-reactive protein and the neutrophil-lymphocyte ratio as a measure of systemic inflammation and with **(C)** the levels of lymphocytes, monocytes and neutrophils. Black lines indicate medians. Differences were assessed using Kruskal -Wallis with Dunn’s multiple comparisons test and considered statistically significant at p<0.05. ** p<0.01, *** p<0.001, **** p<0.0001. The dashed lines indicate the normal thresholds or range values. **(D)** Correlation matrix summarizing the interrelationship observed between the clinical parameters, inflammation markers, blood corpuscles and data from systemic antibody levels measured during acute disease as indicated. The P and R values (Spearman) are shown separately in the mirrored halves of the matrix and have been color-coded as indicated.

**Figure 3.**
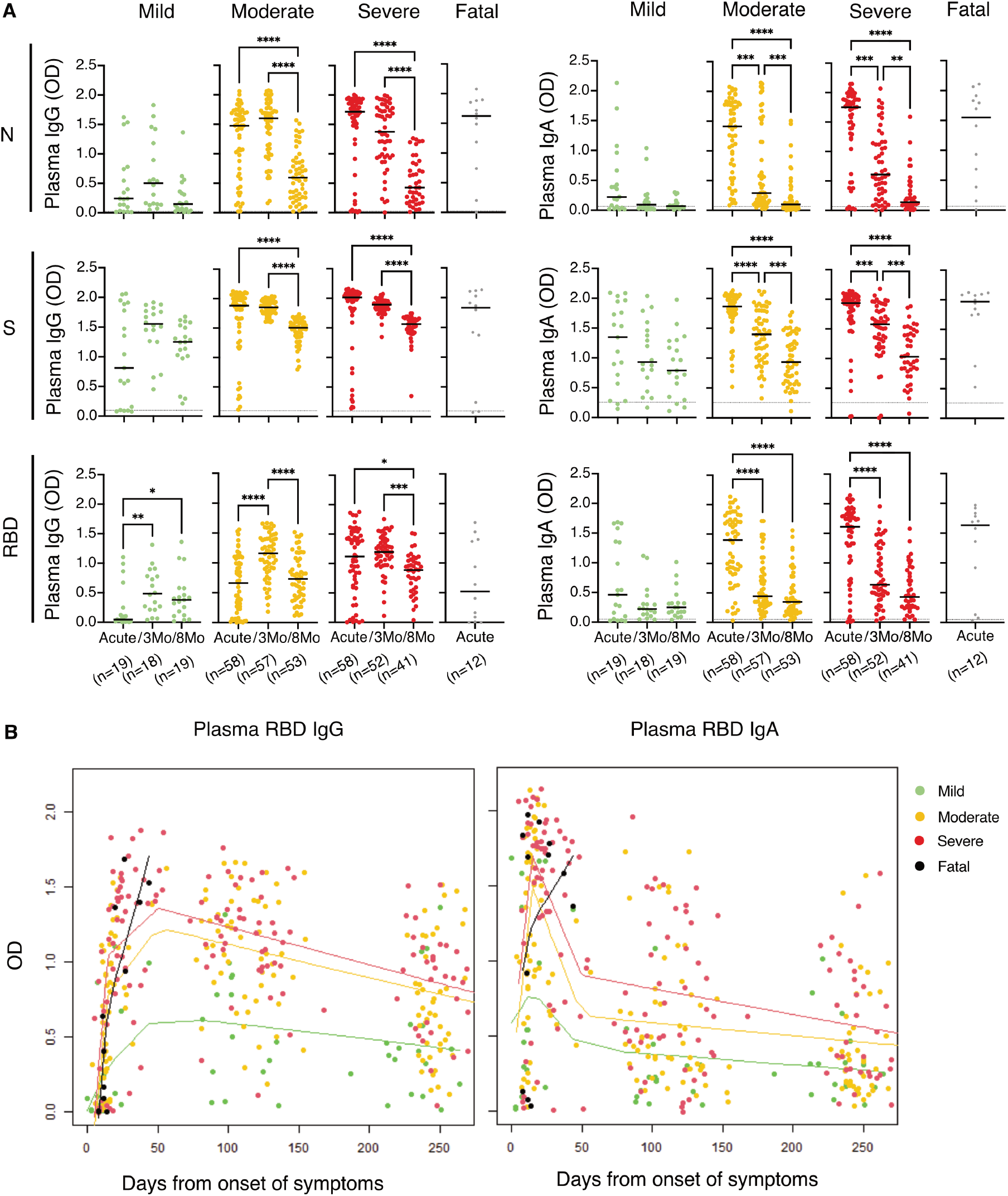
Longitudinal systemic antibody responses across COVID-19 severity from acute disease up to 8 months from symptom onset. **(A)** Individual levels of plasma IgG and IgA (from left to right) in SARS-CoV-2 infected individuals (n=147) with different peak disease severity (PDS). Black lines indicate medians and dotted lines indicate the average background level from pre-pandemic healthy controls. Kruskal-Wallis with Dunn’s multiple comparisons was used to compare the groups and considered statistically significant at p<0.05. ** p<0.01, *** p<0.001, **** p<0.0001. **(B)** Splines graphs of the plasma RBD IgG and IgA level changes over time (n=19 for mild, n=58 for moderate, n=58 for severe and n=12 for fatal). All observations are graphed together with kernel smoothed curves and data points for each group color-coded as previously with the exception of the “Fatal” group which in this figure is highlighted in black. The bandwidth for the smoothing was set to 40, except for the “Fatal” group, for which, due to few and concentrated observations, the bandwidth was set to 10.

The IgG levels had significantly waned from 3 to 8 months in patients who recovered from moderate and severe disease, but the decline was smaller in patients who experienced a mild disease (Figure 3B, Supplementary figure 3 and Supplementary table 1). In contrast to IgG, IgA levels from the acute phase, against all antigens, waned substantially in most patients already after 3 months (Figure 3, Supplementary figure 3 and Supplementary table 1). Antibody titers during acute disease correlated with peak disease severity as well as with disease severity at time of sampling (Supplementary figure 4). The correlation between antibody titers and peak disease severity was maintained also when analyzing the antibodies at the 3- and 8-month follow-up visits (Supplementary figure 4) as also observed in another study (17). Two multivariable linear regression models were also used to estimate the effect of disease severity, days from onset of symptoms, age, gender and CCI on the different plasma antibody levels during the acute phase. One unadjusted model and one model adjusted for these parameters were used (Supplementary table 2). The results from these analyses confirmed the relation between antibody titers and severity as well as the relation between antibodies and days from onset of symptoms (Supplementary table 2).

### Airway IgG and IgA responses and assessment of B cell frequencies in the respiratory tract

We next measured the levels of IgG and IgA in the upper and lower airways and compared with levels in plasma at matched time points. Due to limited respiratory sample volumes, we focused our analyses on IgG and IgA responses against the RBD since these responses are most critical for virus neutralization. We found that RBD-specific antibodies could be detected in nasal swabs (NSW) (Figure 4A) and nasopharyngeal aspirates (NPA) (Figure 4B) during the acute phase across all disease severities (Figure 4A, B and C). In agreement with our observations in plasma, antibody levels in the upper respiratory tract were higher in patients with moderate or severe disease as compared with individuals with mild disease. Both IgG and IgA levels had declined significantly already after 3 months, with IgG declining to almost undetectable levels (Figure 4A-C). RBD antibody levels during acute infection were on average higher in NPA compared to NSW for both IgG and IgA across disease severity (Figure 4A-C) suggesting that antibody titers may increase not only with disease severity but also with sampling at different depths of the upper airways. To address this, we compared the antibody content between donor-matched NSW (peripheral nostril), NPA (upper airway) and ETA (trachea) collected at the same time point during acute disease from intubated patients from whom we had both peripheral, upper and lower airway samples. Interestingly, we still found significantly higher levels of IgA against the RBD in NPA as compared with NSW and ETA (Figure 4D). Furthermore, nasopharyngeal antibody levels (both IgG and IgA) showed a strong correlation with plasma antibody responses (Figure 4E). We also assessed the presence of B cells in the respiratory tract of COVID-19 patients by analyzing the lymphocytes that could be retrieved from NPA and ETA as compared with NPA from three healthy controls (HC). Despite generally obtaining a significantly lower cell yield from NPA as compared with ETA, lymphocyte frequencies did not differ in NPA and ETA from COVID-19 patients but both were lower as compared with NPA from HC. Instead, the proportion of B cells in NPA was higher as compared with ETA in COVID-19 patients and similar to NPA from HC (Figure 5A-B).

**Figure 4.**
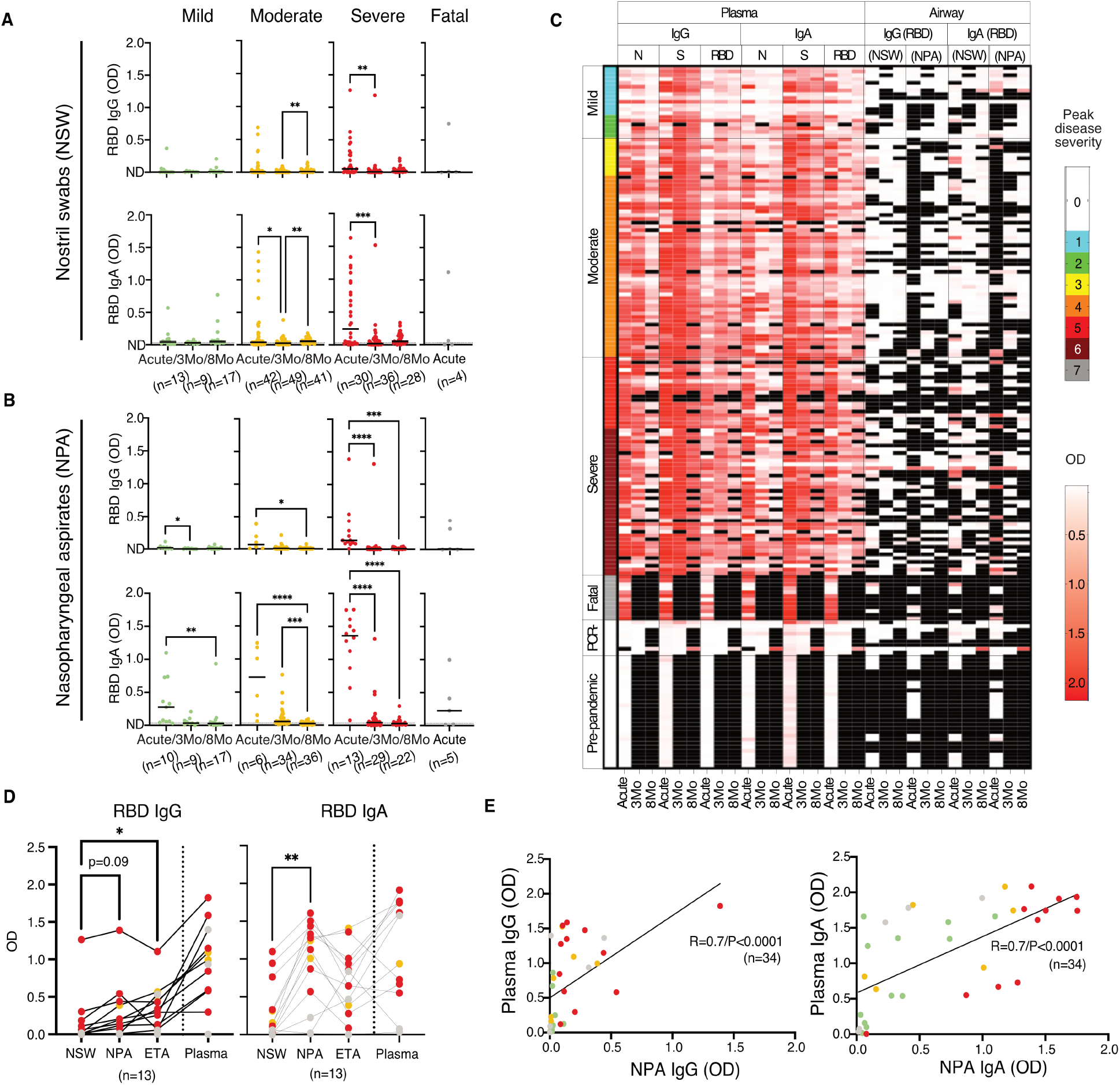
Longitudinal airway antibody responses to RBD across COVID-19 severity from acute disease up to 8 months from symptom onset. Levels of IgG and IgA to RBD in **(A)** nostrils swabs (NSW) and **(B)** nasopharyngeal aspirates (NPA). The black lines indicate median values. Kruskal-Wallis with Dunn’s multiple comparisons was used to compare the groups and considered statistically significant at p<0.05. *p<0.05 ** p<0.01, *** p<0.001, **** p<0.0001. In A) the line overlaps with not detected (ND) for IgG levels. **(C)** Heat map generated grouping patients according to PDS showing acute and convalescent IgG and IgA titers against N, S and RBD (plasma) and RBC (NSW, NPA and ETA) for each patient. The heat map includes data from patients (n=147) and also data from PPHC (n=30) and PCR-individuals (n=9) (indicated with PDS 0). Missing data and not available samples are shown in black. **(D)** Comparison of the levels of RBD IgG/A in patient-matched NSW, NPA, endotracheal aspirates (ETA) and plasma collected at the same time point. The black lines connect data points from the same individuals. Friedman test with Dunn’s multiple comparisons test was used to compare the groups and considered statistically significant at p<0.05. **** p<0.0001. **(E)** Spearman correlation for NPA (n=34) versus plasma immunoglobulins against the RBD during acute disease.

**Figure 5.**
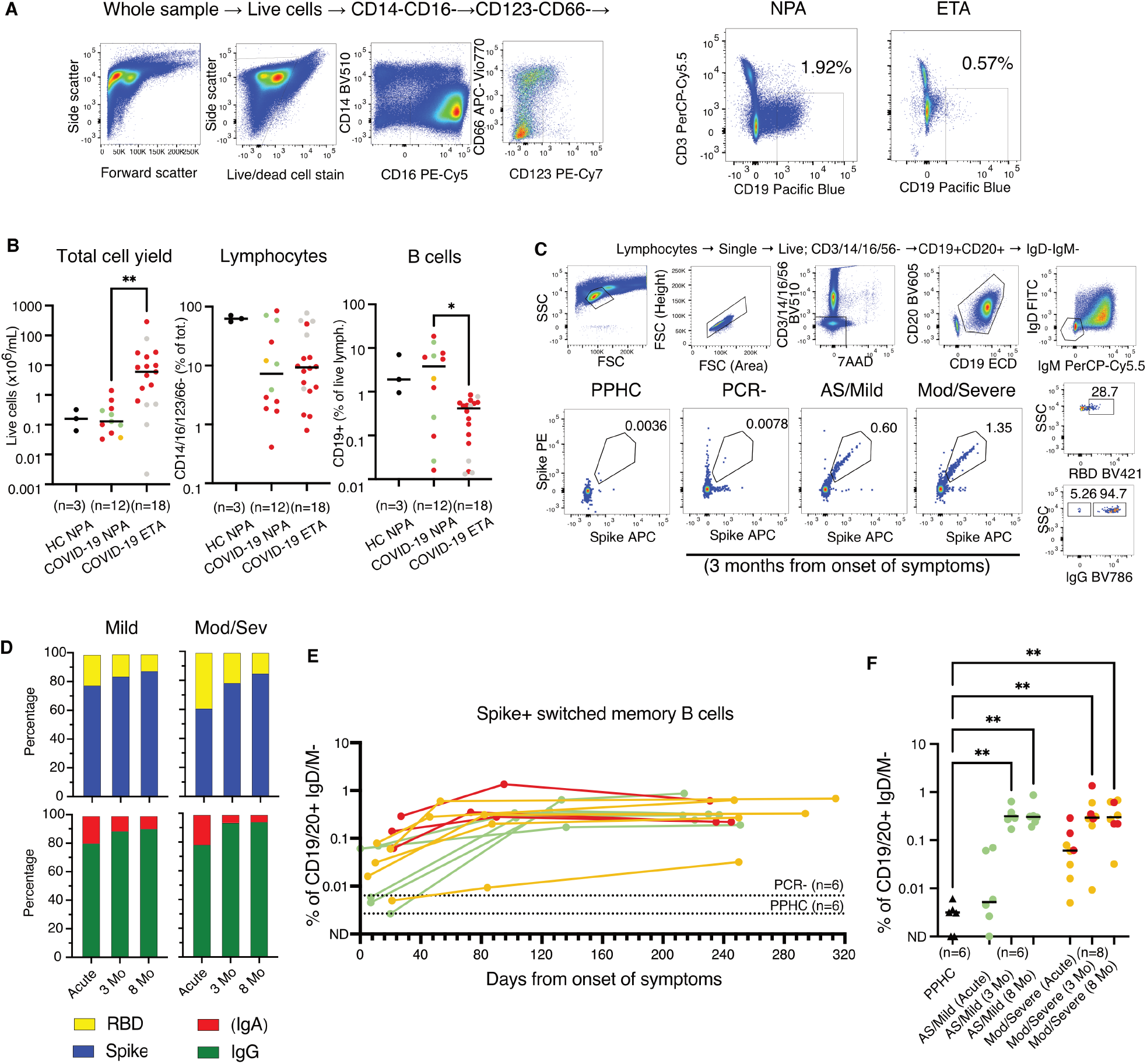
Assessment of frequencies of B cells in the respiratory tract and of circulating S-specific memory B cells. **(A)** Representative example with gating strategy for the identification of lymphocytes (identified as negative for CD14/16/123/66) and of total B cells (CD3-CD19+) in respiratory NPA and ETA samples. **(B)** Lymphocytes and total B cells in NPA and ETA in a subset of patients alongside with NPA from healthy controls. Kruskal-Wallis with Dunn’s multiple comparisons test was used and considered statistically significant at p<0.05. ** p<0.01. **(C)** Representative examples with gating strategy of SARS-Cov-2 S-specific memory B cells from one pre-pandemic healthy control, 3-month follow-up samples from one SARS-CoV-2 PCR-individual and one mild and one moderate/severe COVID-19 patient. Further characterization of S-positive memory B cells on RBD binding and B cell isotype (IgG+ or IgA+ assumed to correspond to IgD-IgM-IgG-B cells). **(D)** Bar charts show the cumulative proportion (frequency) of Spike (blue) and RBD (yellow) specific memory B cell as well as the proportion of IgG (green) vs. IgA (red) isotypes among the Spike specific memory B cells in longitudinal samples from mild (n=6) and moderate/severe (n=8) COVID-19 patients. **(E)** Frequencies of S-specific memory B cells in matched acute (filled) and 3-month follow-up (filled with black lining) PBMCs in relation to days in the subset of individuals analyzed (n=14) color-coded according to PDS. Dotted lines on indicate the average background staining from PCR- and PPHC. **(F)** Levels of circulating Spike+ switched memory B cells during acute disease and convalesce in the subset of patients analyzed, as well as PPHC, color-coded according to PDS. Circles with black lining refer to data during the convalescent phase. Black triangles symbolize the PPHC. Differences were assessed using Kruskal -Wallis with Dunn’s multiple comparisons test and considered statistically significant at p<0.05. ** p<0.01.

### Expansion of SARS-CoV-2-specific memory B cells

As mentioned above, the virus-specific B cell memory pool will be essential to remount a rapid antibody response in the case of re-exposure. To assess the establishment of antigen-specific memory B cells, donor-matched PBMC from acute disease and convalescence were analyzed side-by-side using fluorescently labelled S and RBD probes (18-20). Patients with moderate/severe disease showed the presence of Ig-switched memory B cells specific to S in the acute phase and the memory B cell pool had further expanded after 3 months (ranging from 0.009 to 1.35%; mean 0.42% during convalescence) (Figure 5C-F). Individuals with mild disease showed lower frequencies of S-specific memory B cells during acute disease than the patients with moderate/severe disease. In fact, the frequencies of S-specific memory B cells in the mild patients during the acute phase were not different from those observed in the PCR-individuals or in the PPHC (Figure 5C and E). However, the frequencies of S-specific memory B cells had substantially increased in the mild patients after 3 months (ranging from 0.17% to 0.64%; mean 0.35% during convalescence) and were comparable to frequencies among severe patients. In addition, the levels were well maintained between 3 and 8 months in all groups (Figure 5E and F). Further phenotyping of the S-specific memory B cells indicated that the majority of these cells may be specific for epitopes on S outside of the RBD (Figure 5D). S-specific memory B cells in the circulation were predominantly IgG+, rather than IgA+ (Figure 5D).

### The effect of vaccination on systemic and airway antibody levels

We finally evaluated the influence of SARS-CoV-2 vaccination on the systemic and airway antibody responses (Figure 6A). A subset of 20 individuals, 3 that recovered from mild, 9 from moderate and 8 from severe COVID-19 one year earlier, were sampled after receiving their scheduled vaccination (range 270-407 days; median 339 days from symptom onset) (Table 2). Donor-matched plasma, NSW and NPA were collected at different timepoints after prime (7-16 days) from 18 patients and after boost (7-28 days) from 19 patients alongside with samples from 12 individuals naïve to SARS-CoV-2 (7-10 days after prime and boost vaccinations) to be included as a reference control. All samples were analyzed for the presence of IgG and IgA against RBD. Antibodies against N were also measured in patient plasma as a negative control as the vaccines used were based on the S protein. After vaccination, all individuals demonstrated a significant increase of both plasma IgG and IgA against the RBD (Figure 6B) but, as expected, not against N (Figure 6B). While the IgG levels to RBD after boost vaccination exceeded the levels detected during the acute phase, the IgA levels were equally high (Figure 6B). On the contrary, individuals naïve to SARS-CoV-2 only had a moderate increase of IgA as compared with IgG after boost (Figure 6B). IgG levels after boost were significantly lower in individuals naïve to SARS-CoV-2 as compared with those from COVID-19 patients after boost (Figure 6C and Supplementary figure 5A). The airway IgG levels to RBD also showed a noticeable increase after the boost vaccination in particular. In fact, the IgG levels in the airway samples, both nasal swabs and NPA, were in many individuals significantly higher after boost vaccination than they were in the acute stage of the disease (Figure 6D). In contrast, this was not noted for IgA levels to RBD (Figure 6D). On the other hand, individuals naïve to SARS-CoV-2 only had a modest but significant increase of IgG in NSW and NPA and of IgA in NSW after boost (Figure 6D). Despite IgG levels in NSW had the highest increase after boost in individuals naïve to SARS-CoV-2, levels were generally significantly lower as compared with those from COVID-19 patients (Figure 6E and Supplementary figure 5A).

**Figure 6.**
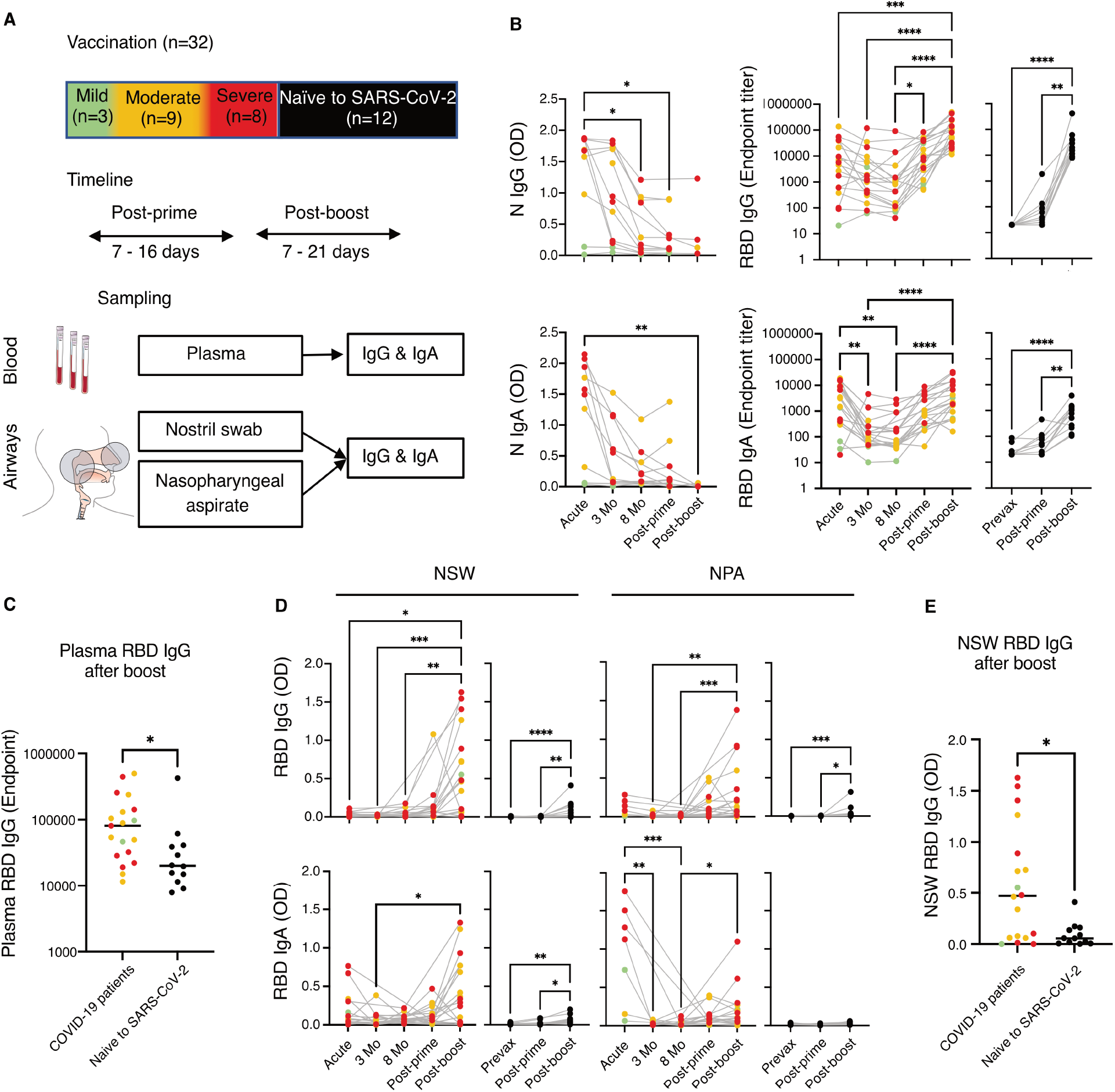
Vaccination and systemic and airway antibody level rebound. **(A)** Overview of vaccinated patients (n=20) with respect with peak disease severity during COVID-19 and sampling timeline after prime and boost as compared with vaccination in individuals naïve to SARS-CoV-2 (n=12). The anatomical compartments analyzed and the measurements performed are also shown. **(B)** Compiled patient-matched longitudinal data from acute, 3-month- and 8-month follow-ups are shown together with data from after prime and after boost for the levels of plasma IgG and IgA against N and RBD. **(C)** Direct comparison between plasma RBD IgG after boost in COVID-19 patients and individuals naïve to SARS-CoV-2. **(D)** Compiled data as above for RBD IgG and IgA in NSW and NPA. **(E)** Direct comparison between NSW RBD IgG after boost in COVID-19 patients and individuals naïve to SARS-CoV-2. The grey lines connect data points from the same individuals. Data are color-coded according to peak disease severity during COVID-19 with data from individuals naïve to SARS-CoV-2 shown in black as a comparison. Differences were assessed using Kruskal -Wallis with Dunn’s multiple comparisons test and considered statistically significant at p<0.05. ** p<0.01, *** p<0.001, **** p<0.0001.

**Table 2.**
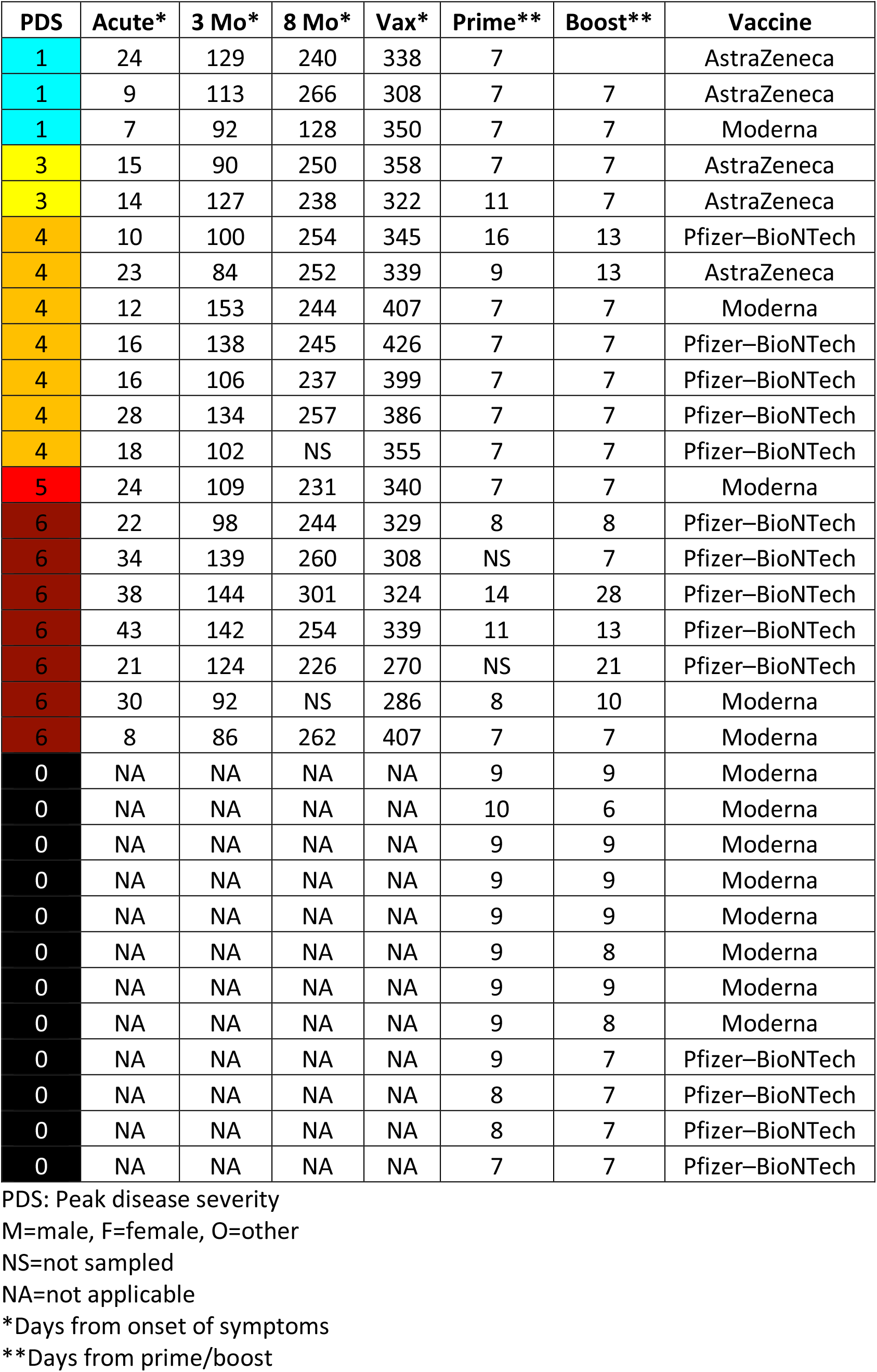
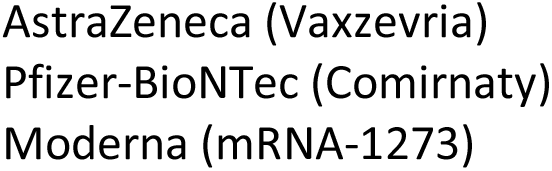
Peak disease severity and longitudinal sampling timeline of patients and individuals naïve to SARS-CoV-2 vaccinated against SARS-CoV-2.

## Discussion

By now, it is well documented that higher systemic antibody levels are generated in severe as compared with mild COVID-19 (7-11, 21-23). In contrast, the presence and durability of antibodies against SARS-CoV-2 in the airways is much less understood. Nor is it known if and how respiratory antibody levels are influenced by SARS-CoV-2 vaccination in humans. In this study, we performed longitudinal analyses of systemic and upper and lower airway antibody responses in a clinically well-characterized and relatively large cohort of individuals with SARS-CoV-2 infection representing the full spectrum of COVID-19 severity ranging from asymptomatic infection to fatal disease. Matched analyses in blood and in the airways enabled us not only to address the magnitude and durability of systemic antibodies to SARS-CoV-2 but also to gain insights into the prospects of protective capacity locally in the mucosa at virus re-entry. This is one key aspect still largely unknown yet critical for our understanding of immunity to and protection from SARS-CoV-2. Furthermore, we studied how the systemic versus airway antibody levels were affected by vaccination. Collectively, this data will contribute to a better understanding of long-term protective effects and whether vaccination is important to boost the capacity of virus neutralization in the airways and thus reducing re-infection and virus spread.

Airway mucus along the respiratory tract is thought to serve as a barrier that can trap respiratory viruses via virus glycoprotein-mucin interactions (24). However, it has been shown that local immobilization of respiratory viruses such as influenza viruses in the airways mostly occurs by binding with virus-specific antibodies present in the mucus (25). As the respiratory tract is the initial site of viral infection and replication, the levels of IgG and IgA against the RBD in the upper and lower airways are likely critical for SARS-CoV-2 neutralization and could therefore help predict the ability of individuals to neutralize the virus locally in case of re-exposure. Low but detectable levels of antibodies to SARS-CoV-2 have previously been reported in saliva during convalescence (26). However, measurements of antibodies in saliva may primarily represent plasma exudate from the gingiva (27) while respiratory secretions better reflect the mucosal responses. Sampling the respiratory mucosa is indeed more likely to be sensitive to sampling methods compared to blood draws. Ideally it is therefore important to sample multiple compartments to more comprehensively understand the immunity to SARS-CoV-2. In our study we found that IgG and IgA against the RBD can be readily detected in the upper and lower airways during acute disease and that such levels correlated with the systemic response at the same time point and also followed disease severity. However, for all the patients across disease severities, airway antibodies waned to low levels much faster than those in plasma during convalescence. Whether these low antibody levels observed at respiratory sites will be sufficient for preventing virus re-entry or for protection is not known. The correlation between systemic and airway antibody levels during acute disease raises questions on whether the low levels of antibodies in the airways during convalescence are due to decreased antibody generation locally at mucosal sites or are rather caused by decreased dissemination from the periphery once systemic antibody levels start to wane. Antibodies in the upper respiratory tract have been shown to be dominated by secretory IgA which are mostly produced by plasma cells in the lamina propria of mucosa-associated lymphoid tissue (MALT) (28, 29). We detected high levels of IgA in the upper airways early during acute COVID-19 that rapidly declined during convalescence, following the pattern observed for systemic IgA levels here and in other reports (30-32). This suggests that at least some IgA disseminated into the airways from the circulation. In contrast, the dynamics of IgG were different in the respiratory samples compared to plasma with airway IgG following the same kinetics as IgA, while systemic IgG were well maintained up to 8 months.

When we assessed the presence of lymphocytes in the different airway compartments during acute disease, we observed higher proportion of B cells along with high antibody levels, especially IgA, in the nasopharynx, as compared with the nostril or the endotracheal space. It has previously been shown that the majority of antibody secreting cells generated after intranasal immunization with live-attenuated vaccines in rodents may reside in the respiratory tract rather than in the spleen and bone marrow (33) and that these cells secrete IgA early after a later challenge with the vaccination pathogen (34-36). Therefore, it is possible that B cells generated during SARS-CoV-2 infection also reside locally in the airways and contribute to antibody levels in the nasopharynx. While the antibody content in NPA and ETA could be influenced by differences in sampling methods and sample volumes, these data suggest that antibody abundance and possibly virus neutralization via IgA differ along the respiratory tract and may be more pronounced in the nasopharynx compared to the lower airways. Altogether, our observations demonstrate that moderate and severe COVID-19 result in high levels of circulating antibodies and despite that IgG levels are well-maintained, antibody levels in the airways decline significantly after the acute phase.

Once antibody titers have waned below protective levels, the response to a secondary infection will mostly rely on the presence of resting antigen-specific memory B cells that can rapidly activate upon antigen re-exposure (13). Therefore, similar to other studies (18-20), we investigated the induction and maintenance of S-specific memory B cells. Importantly, because of the comprehensive range of disease severity represented in our cohort, we were able to compare the opposite ends of the COVID-19 disease spectrum by focusing on individuals with mild disease as compared with patients with moderate/severe disease who had the highest circulating IgG and IgA levels. Strikingly, despite the fact that these patients were at the opposite ends of the disease severity spectrum, they had comparable levels of S-specific memory B cells during convalescence. These appeared to be specific for epitopes on S outside of the RBD and were predominantly IgG+, rather than IgA+, which may affect the proportions of different isotypes subsequently produced in the event of antigen re-exposure.

Immunization at mucosal sites such as for example intranasal administration of live-attenuated influenza vaccines generally elicits mucosal immune responses (37). However, several studies, primarily performed with DNA and virus-like particles (VLP) vaccines, have shown that intradermal, subcutaneous and intramuscular immunization also can result in local mucosal responses that protect from mucosal challenge (38). It has been speculated that this could be due to free antigen or B cells migrating from the vaccine draining lymph nodes to the MALT (38-40). A two-dose regimen of Moderna’s mRNA-1273 vaccine administered intramuscularly and followed by intranasal and intratracheal challenge with SARS-CoV-2 in rhesus macaques has indeed shown to result in local virus neutralization in the airways (41). Antibodies in bronchoalveolar lavage and nasal swabs were elicited in a vaccine dose-dependent manner assessed after the boost vaccination (42).

Whether the systemic and/or mucosal immunity generated during natural infection is boosted by vaccination and results in a similar or enhanced magnitude of responses would be important knowledge to acquire for planning the best vaccination strategies for SARS-CoV-2 as well as for other respiratory viruses. Our results on individuals recovering from COVID-19 and subsequently receiving vaccination indicated a marked increase of both IgG and IgA levels systemically but also strikingly in the airways, which in the majority of cases exceeded the levels observed during acute disease. In contrast, vaccination of individuals naïve to SARS-CoV-2 only resulted in a modest increase of airway antibodies, mainly IgG, after boost vaccination. Notably, the antibody increase observed between prime and boost vaccination in the patients was more prominent in the airways than systemically. Recent studies on systemic antibody responses after SARS-CoV-2 vaccination in individuals who recovered from COVID-19 have shown a significant increase in antibody levels after one vaccine dose with no or only a small increase after the second dose (43-47). This suggests that one vaccine dose may be sufficient to protect these individuals from disease in case of re-infection which is important for vaccine dose management at the population level. However, our data indicate that only assessing the systemic antibody levels after vaccination is to some extent misleading as respiratory antibody levels, and likely virus neutralization, may be substantially better with a prime-boost vaccination strategy rather than with one single dose. Two earlier studies have been able to demonstrate neutralizing activity of antibodies in the upper respiratory tract after vaccination in individuals who earlier had COVID-19 (49, 50).

The higher levels of airway antibodies that we observed after two vaccine doses may be explained by that even a small increase in circulating antibodies after the boost causes a substantial extravasation from the bloodstream into mucosal sites (Supplementary figure 5B). On the other hand, the fact that naïve individuals had less pronounced airway antibodies after vaccination despite they elicited relatively high plasma antibody levels suggests that airway antibody responses are better primed with natural infection and that vaccination after COVID-19 stimulates anamnestic local mucosal responses. It remains to be investigated how airway antibodies induced after intranasal vaccination would compare to natural infection and whether an intramuscular vaccine boost would affect these responses (48).

In summary, here we show that COVID-19 disease severity not only determines the magnitude of systemic but also airway antibody levels with efficient generation of virus-specific memory B cells against SARS-CoV-2 also occurring upon mild disease. While plasma IgG levels were generally well detectable after acute disease in all groups, there was a significant decline in airway antibodies during convalescence. This suggests that antibodies in the airways may not be maintained at levels that prevent local virus entry upon re-exposure. However, our data indicate that the majority of infected individuals have the ability to generate anamnestic responses via the memory B cell pool and that vaccination against SARS-Cov-2 resulted in a substantial rebound of both systemic and airway antibodies in patient who recovered from COVID-19. These data indicate a positive effect of vaccination for increased virus neutralization in the airways and prospects of reduced virus spread, which further supports following the full vaccination schedule also in this population.

## Methods

### Study design, patient enrollment and sample collection

One hundred and forty-seven (147) PCR-confirmed SARS-CoV-2 infected patients were enrolled at the Karolinska University Hospital and Haga Outpatient Clinic (Haga Närakut), Stockholm, Sweden during March-May 2020 (acute phase) in a time that ranged from 0 to 54 days from onset of symptoms as self-reported by individual patients; and during April-September 2020 (3 months) in a time that ranged from 46 to 168 days and during November 2020 to February 2021 (8 months) continuing from the previous counts. Patients were enrolled at various settings, ranging from primary to intensive care. In order to recruit asymptomatic and mild cases, household contacts of COVID-19 patients were enrolled and screened with PCR to identify SARS-CoV-2 positive individuals. A small subset of these individuals who experienced influenza-like symptoms and were possibly exposed to SARS-Cov-2 but had a negative diagnostic PCR (PCR-) (n=9 of whom 3 were household contacts of confirmed patients with 1 experiencing fever, and 6 were included based on suspected infection with 4 experiencing fever) were sampled in the same way and included as controls alongside with 30 pre-pandemic healthy control samples (PPHC) from 2016-2018. Twelve individuals naïve to SARS-CoV-2 were also recruited at the Umeå University, Umeå, Sweden as a control group for vaccination (Table 2). These individuals were identified as naïve to SARS-CoV-2 based on lack of COVID-19 symptoms and positive diagnostic PCR test throughout the pandemic in combination with absence of plasma antibodies against SARS-CoV-2 prior to vaccination.

Respiratory failure was categorized daily according to the respiratory domain of the Sequential Organ Failure Assessment score (SOFA) (14). The modified SOFA score (mSOFA) was calculated when arterial partial pressure of oxygen (PaO_2_) was not available. In this case peripheral transcutaneous hemoglobin saturation (SpO_2_) was used instead (15). Estimation of the fraction of inspired oxygen (FiO_2_) based on O_2_ flow was calculated as per the Swedish Intensive Care register definition (51). Patients were categorized based on the peak respiratory SOFA or mSOFA value with the 4-point respiratory SOFA score being extended with additional levels to distinguish between admitted and non-admitted mild cases (both respiratory SOFA score 0) and to include fatal outcome. Ten (10) patients with fatal outcome had peak disease severity score 6 prior to death and 2 patients had scores of 4 and 5. For convenience, the resulting 7-point composite peak disease severity (PDS) was condensed into a broader classification consisting of mild (1-2), moderate (3-4), severe (5-6), and fatal (7). Demographics and additional data were collected from medical records, including clinical history and risk factors such as BMI and co-morbidities. Total burden of comorbidities was assessed using the Charlson co-morbidity index (CCI) (52) (Table 1). Additional clinical information on this patient cohort including the modulation of disease from time to study inclusion to peak severity can be found in Falck-Jones et al (16).

Blood was collected in EDTA-containing tubes from all patients except those admitted to the intensive care unit (ICU) for whom blood was pooled from heparin-coated blood gas syringes discarded in the last 12 hours. For some ICU patients, additional venous blood was also collected in EDTA tubes. Nostril swabs (NSW) and nasopharyngeal aspirates (NPA) were collected from the majority of the patients whereas endotracheal aspirates (ETA) were only collected from patients with mechanical ventilation intubated in the ICU. Admitted patients were sampled during acute disease at up to four timepoints and ICU patient material was collected up to ten timepoints. For this study, unless otherwise stated, the measurements referring to acute disease were performed with samples collected at the time of study inclusion and when patients returned for their follow-up visits at 3 and 8 months from symptom onset. At follow-up sampling, all study individuals had been discharged (if hospitalized) from the infectious diseases ward but some individuals (<10) who recovered from severe COVID-19 were still in a hospital aftercare ward at the first follow-up sampling. All study participants were confirmed SARS-CoV-2 negative by PCR at the time of follow-up sampling, with the exception of 5 individuals who were PCR+ but with high Ct values (>34).

### Enzyme-linked immunosorbent assay (ELISA)

The presence of IgG or IgA binding against the SARS-CoV-2 Nucleocapsid (N) and Spike (S) trimer or the Receptor Binding Domain (RBD) monomer (5, 6) in plasma and airway samples was assessed by enzyme-linked immunosorbent assay (ELISA). Recombinant proteins were received through the global health-vaccine accelerator platforms (GH-VAP) funded by the Bill & Melinda Gates Foundation, Seattle, WA, USA. Briefly, 96-half well plates were coated with 50ng/well of the respective protein. Plates were incubated with a selected duplicate dilution that did not provide background noise against ovalbumin used as a negative control (data not shown) (i.e. 1:20 for plasma samples, 1:2 for NSW and NPA, and 1:5 for ETA in 5% milk/PBS buffer). Duplicate 7-point serial dilutions were initially performed for measuring plasma IgG against RBD during acute disease and after vaccination. The half maximal effective concentration (EC_50_) or the endpoint titer (dilution at the set OD value of 0.1) were calculated using GraphPad Prism 9. The relation between EC50 and endpoint titer for these samples is shown in Supplementary figure 5C. However, since for several samples with low antibody concentration (mostly from the asymptomatic/mild category) the EC50 was below the highest dilution used (of 1:20) and therefore below the limit of detection (Supplementary figure 6A), the maximal optical density (OD) at 1:20 dilution was used for most of the analyses. The relation between maximal OD and EC50 was verified in a subset of patients with high IgG and IgA against S (Supplementary figure 6B). To be able to compare pre- and post-vaccination antibody levels that would, in some instances, fall below and above the lower and upper limits of detection, the endpoint titer was used instead.

Detection was performed with mouse and goat anti-human IgG or IgA HRP-conjugated secondary antibodies (clone G18-145 from BD Biosciences and polyclonal from ThermoFisher, respectively) followed by incubation with TMB substrate (BioLegend) which was stopped with a 1M solution of sulfuric acid. Blocking with 5% milk/PBS buffer and washing with 0.1% Tween-20/PBS buffer were performed between each step. Absorbance was read at 450nm and background correction at 550nm using an ELISA reader. Data were reported as maximal absorbance i.e. OD, as stated above, and plotted using GraphPad Prism 9. All of the antibody measurements in plasma and respiratory samples from SARS-CoV-2 patients were run alongside with samples from two different control groups as described above. Interestingly, low but readily detectable IgA reactivity against S was detected in the pre-pandemic healthy controls and in the PCR-individuals (Supplementary figure 6C). After having verified the specificity and sensitivity of our ELISA assay for IgA detection with limiting sample dilutions (Supplementary figure 6D-E), we hypothesize that this might be due to cross-reactivity on the shared portions of the S protein between SARS-CoV-2 and other common cold coronaviruses. Reports have shown that cross-reactivity between coronaviruses exists (53, 54).

### Flow cytometry

Staining of cells from airway samples was performed fresh. Briefly, samples were centrifuged at 400 g for 5 min at room temperature and cells were washed with sterile PBS. Mucus was removed using a 70 μm cell strainer and cells were subsequently stained with the appropriate combination of fluorescently labelled monoclonal antibodies as illustrated in Figure 5A and in Table 3A. Staining of PBMC was performed on previously cryopreserved samples. The appropriate combination of fluorescently labelled monoclonal antibodies binding to different cell surface markers and with fluorescently labelled S and RBD proteins used as probes for antigen-specific B cells is illustrated in Figure 5C and in Table 3B. Probes were prepared from biotinylated proteins using a 4:1 molar ratio (protein:fluorochrome-labelled streptavidin) considering the molecular weight of protein monomers and of the streptavidin only. The probes were prepared using streptavidin conjugated to PE and APC for S and with BV421 for the RBD. The gating strategy for the identification of antigen-specific memory B cells is shown in Figure 5C. Briefly, after identification of lymphocytes in single suspension, live B cells, (i.e. cells not expressing CD3/,CD14/CD16/CD56) were gated. From this gate, B cells were further isolated by expression of CD19 and CD20 and then switched memory B cells were identified as IgD-IgM-. From these, S-specific switched memory B cells were identified by binding to both S protein probes. Further characterization was then carried out by analyzing IgG expression (IgA+ switched memory B cells are assumed to mirror IgD-IgM-IgG-B cells) and fluorescently labelled RBD. Stained cells from airway samples were acquired using a BD LSRFortessa while stained PBMC were acquired using a BD FACSAria Fusion both interfaced with the BD FACSDiva Software. Results were analyzed using BD FlowJo version 10.

**Table 3.**
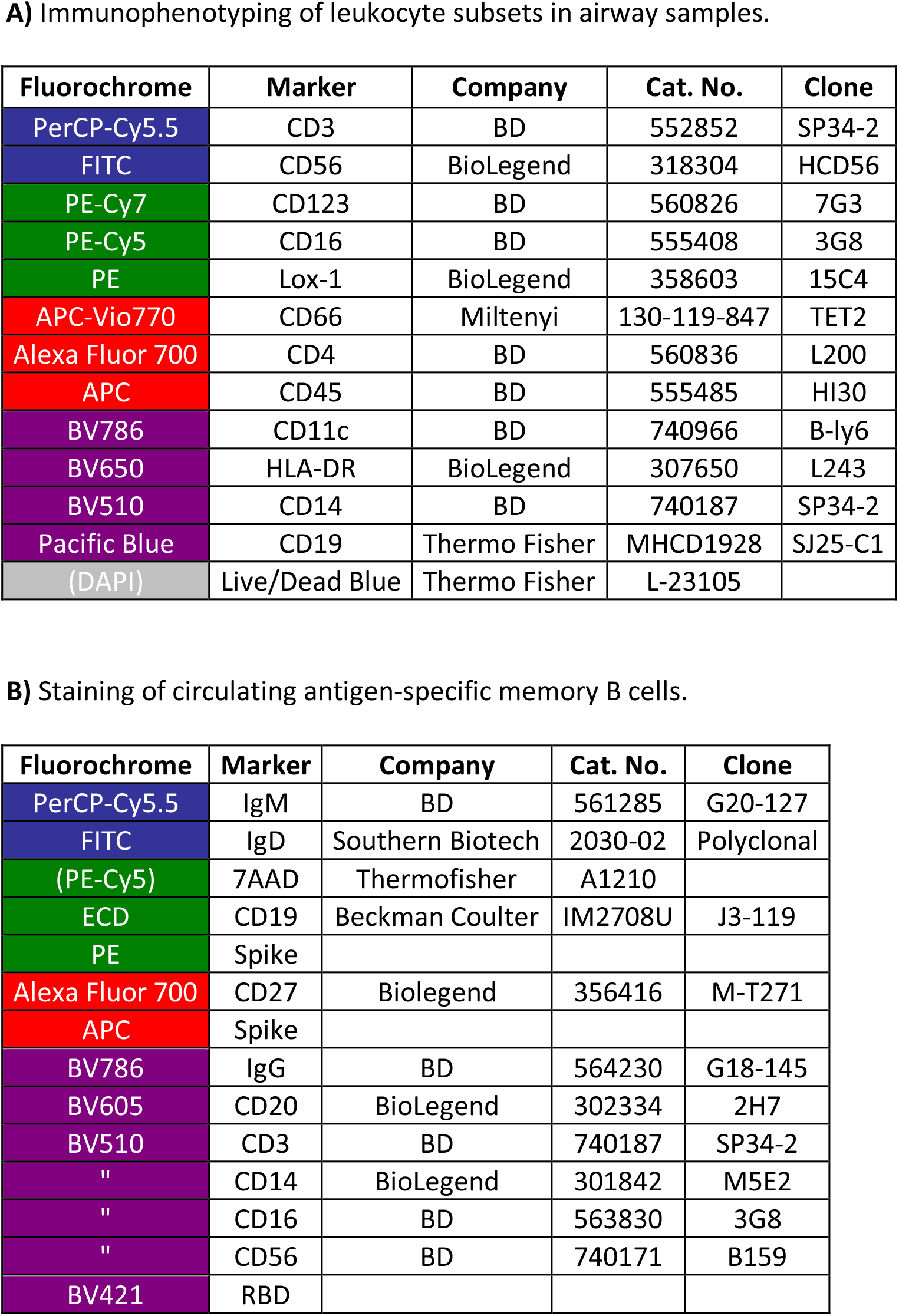
Flow cytometry panels.

### Statistics

Spearman correlation was used to assess the interdependence of 2 different non-categorical parameters across individuals whereas Wilcoxon matched-pairs signed rank or Mann– Whitney U tests as appropriate, were used to assess differences or similarities for one single parameter between 2 different groups. Kruskal -Wallis with Dunn’s multiple comparisons test was used when assessing comparison between multiple groups. All of the above statistical analyses were performed using GraphPad Prism 9.

The effect of disease severity on the acute response was estimated using linear regression. We estimated both unadjusted models, as well as models adjusted for age, gender, days from onset of symptoms and CCI (Charlson Comorbidity Index). The longitudinal models using splines were estimated using multivariate multiple regression. The splines used were linear, with knots placed on days 15 and 50. The location of the knots was chosen based on visual inspection of the data, aided by kernel smoothing. The effect on standard deviations from repeated measures was not adjusted for, as the primary focus of the longitudinal analysis was description rather than statistical testing. Analysis was done in R, version 4.1.0 (R Core Team (2021). R: A language and environment for statistical computing. R Foundation for Statistical Computing, Vienna, Austria. URL https://www.R-project.org/.)

When not stated otherwise, p-values smaller than 0.05 were considered statistically significant.

### Study approval

The study was approved by the Swedish Ethical Review Authority, and performed according to the Declaration of Helsinki. Written informed consent was obtained from all patients and controls. For sedated patients, the denoted primary contact was contacted and asked about the presumed will of the patient and to give initial oral and subsequently signed written consent. When applicable, retrospective written consent was obtained from patients with non-fatal outcomes.

## Data Availability

The data that support the findings os this study are available from the corresponding author on reasonable request.

## Author contributions

Experimental study design: A.C., Ka.L. and A.S-S. Clinical concept design: A.S-S., M.B., N.J., J.A., J.S., A.F. and M.F. Acquisition and sample processing: A.C., M.Y., S. F-J., S.V., B.Ö., E.Å., L.A., R.F-J., M.Ö., F.G., J.S., M. E. J.M. and C.A. Generation of data: A.C., M.Y., S. F-J., S.V., L.A. and P.C.G. Provision of custom reagents: D.P., M.M., L.C., and N.P.K. Analysis and interpretation of data: A.C., Ka.L. and A.S-S. Critical revision of the manuscript: all authors. Statistical analysis: A.C., A.W., K.L. and S.O. Ka.L. and A.S-S. contributed equally to the study.

## Acknowledgments

We thank the patients and healthy volunteers who have contributed to this study. We would also like to thank Alicia Edin, Andy Dernstedt, Mikaela Lagerkvist, Emma Stenlund, Maj Järner, medical students and hospital staff for assistance with patient sampling, collection of clinical data and sample processing, the Biomedicum BSL3 core facility, Karolinska Institutet and Fredrika Hellgren for assistance with English editing. This work was supported by grants from the Swedish Research Council, the Swedish Heart-Lung Foundation, the Bill & Melinda Gates Foundation, the Knut and Alice Wallenberg Foundation through SciLifeLab and Karolinska Institutet.

## Notes

**Conflict of interest statement** The authors have declared that no conflict of interest exists.

### Competing Interest Statement

The authors have declared no competing interest.

### Author Declarations

The study was approved by the Swedish Ethical Review Authority, reference: 2015/1949-31/4 (COVID-19 patients) and reference: 2021-00055 (SARS-CoV-2 naive individuals), and performed according to the Declaration of Helsinki.

### Summary of Updates

We have now performed more advanced statistics on the analysis of longitudinal data and included data on respiratory antibody responses after vaccination of SARS-CoV-2 naive individuals.

