## Supplemental data for "Airway antibodies emerge according to COVID-19 severity and wane rapidly but reappear after SARS-CoV-2 vaccination"

[1]Division of Immunology and Allergy, Department of Medicine Solna, Karolinska Institutet, Karolinska University Hospital, Stockholm, Sweden. [2]Division of Biostatistics, Institute of Environmental Medicine, Karolinska Institutet, Stockholm, Sweden. [3]Department of Physiology and Pharmacology, Karolinska Institutet, Stockholm, Sweden. [4]Department of Perioperative Medicine and Intensive Care, Karolinska University Hospital, Stockholm, Sweden. [5]Division of Infectious Diseases, Department of Medicine Solna, Center for Molecular Medicine, Karolinska Institutet, Sweden. [6]Department of Infectious Diseases, Karolinska University Hospital Solna, Stockholm, Sweden. [7]Department of Microbiology, Tumor and Cell Biology, Karolinska Institutet, Stockholm, Sweden. [8]Clinical Microbiology, Karolinska University Hospital Solna, Stockholm, Sweden. [9]Närakut SLSO, Karolinska University Hospital Solna, Stockholm, Sweden. [10]Department of Biochemistry, University of Washington, Seattle, WA, United States. [11]Institute for Protein Design, University of Washington, Seattle, WA, United States. [12]Section of Infection and Immunology, Department of Clinical Microbiology, Umeå university, Umeå, Sweden. [*]Equal contribution.

**Correspondence to:** Karin Loré and Anna Smed-Sörensen, Division of Immunology and Allergy, Department of Medicine Solna, Karolinska Institutet, Visionsgatan 4, BioClinicum J7:30, Karolinska University Hospital, 171 64 Stockholm, Sweden.

**Conflict of interest statement**

The authors have declared that no conflict of interest exists.

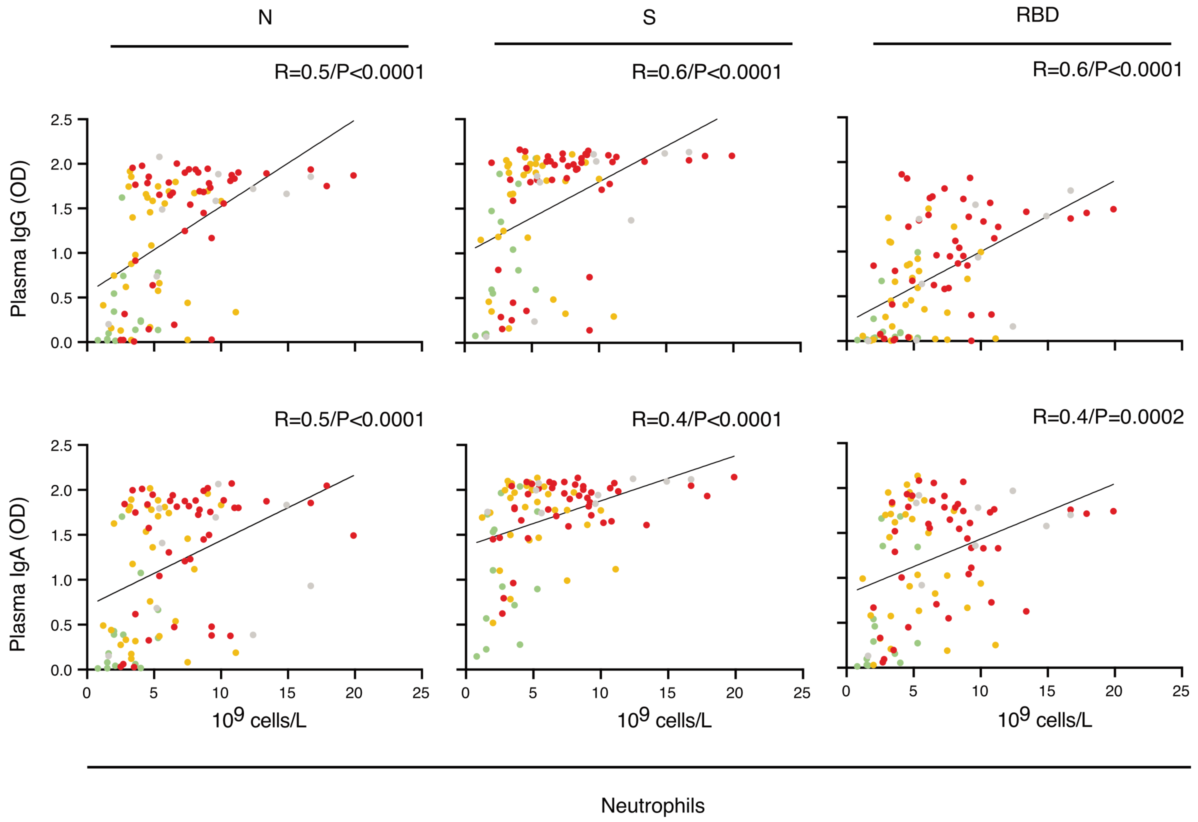


**Supplementary figure 1.** Spearman correlation for plasma IgG (top) and IgA (bottom) against the N, S and RBD versus the level of neutrophils during the acute phase (n=147). Data in cyan and green refer to mild disease (PDS 1 and 2), yellow and orange refer to moderate disease (PDS 3 and 4), red and cayenne refer to severe disease (PDS 5 and 6) and grey refers to patients with fatal outcome (PDS 7).

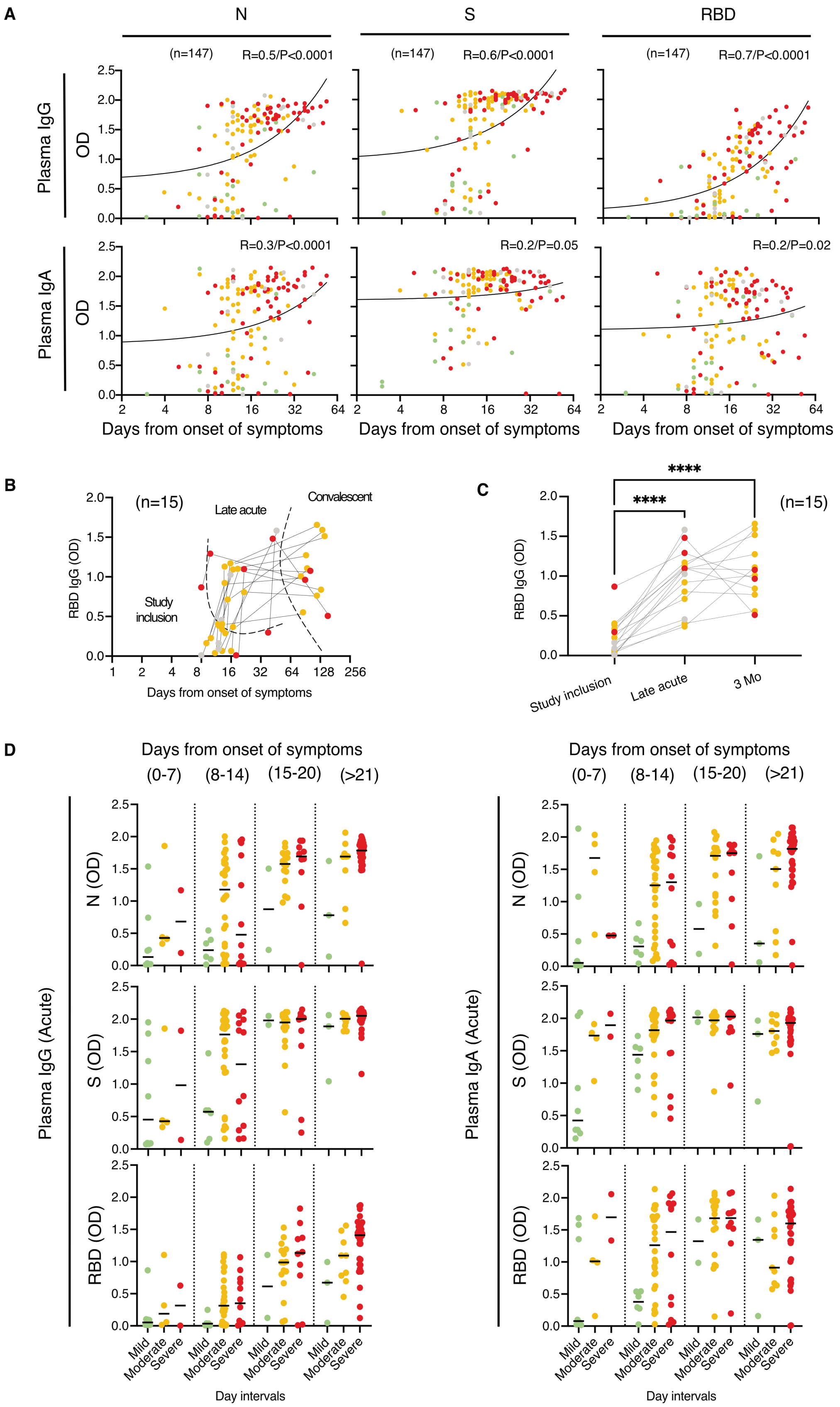


**Supplementary figure 2. (A)** Spearman correlation for plasma immunoglobulins against the N, S and RBD versus days from onset of symptoms during the acute phase are shown (n=147). **(B-C)** Longitudinal measurements of plasma IgG against RBD on a subset of patients with moderate/severe disease and fatal outcome who had low (< 1.0 OD) antibody titers at the time of study inclusion when longitudinal samples from symptomatic (acute) disease were available. Levels are shown at the time of study inclusion (8-38 days after onset of symptoms), during the late acute phase (10-47 days after onset of symptoms) and at convalescence (3 months), and shown with respect to days from onset of symptoms (the different time intervals are separated by dotted lines) and as a group comparison. The black lines connect data points from the same individuals. Kruskal-Wallis with Dunn's multiple comparisons test was used to compare the groups separately and considered statistically significant at $p<0.05$. **** $p<0.0001$. **(D)** Analysis on IgG and IgA against N, S and RBD during the acute phase (n=147), in patients grouped with respect to days from onset of symptoms. Differences within each time interval, were assessed using Kruskal-Wallis with Dunn's multiple comparisons test. No significant differences were found. Single data points in cyan and green refer to mild disease with peak disease severity (PDS) 1 and 2, data in yellow and orange refer to moderate disease (PDS 3 and 4), data in red and cayenne refer to severe disease (PDS 5 and 6) whereas data in grey refer to patients who had a fatal outcome (PDS 7).

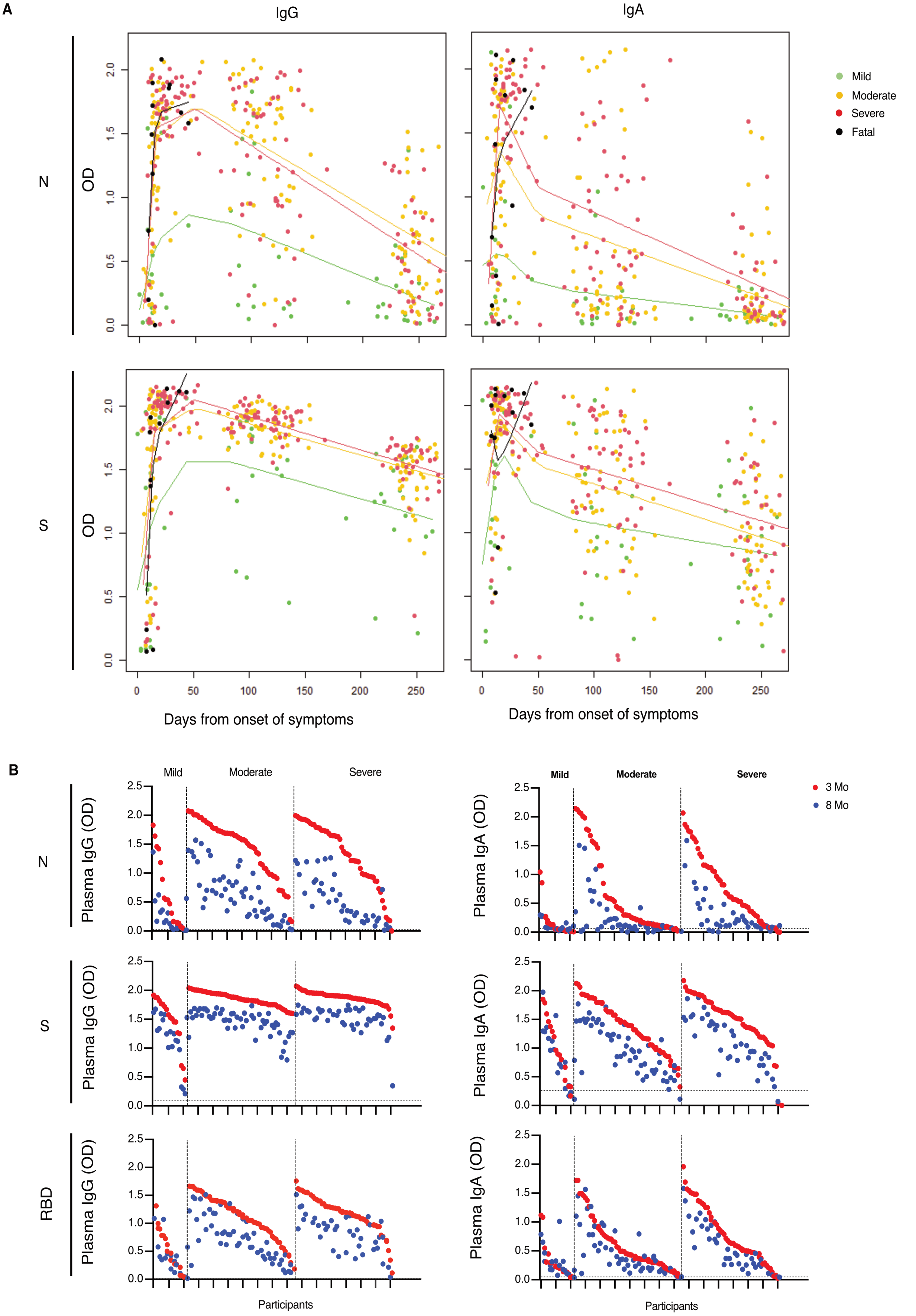


**Supplementary figure 3. (A)** Spline graphs of the plasma N and S IgG and IgA level changes over time (n=147). All observations are graphed together with curves and data pointes for each group color-coded as previously with the exception of the "Fatal" group which in this figure is highlighted in black. The bandwidth for the smoothing was set to 40, except for the "Fatal" group, for which, due to few and concentrated observations, the bandwidth was set to 10. **(B)** Matched individual levels of plasma IgG and IgA measured during convalescence at the 3-month follow up-visit (n=128) (in red) and later after 8 months (n=113) (in blue). Data sets for each measurement have been ordered according with decreasing titers observed at 3 months individually for each disease severity.

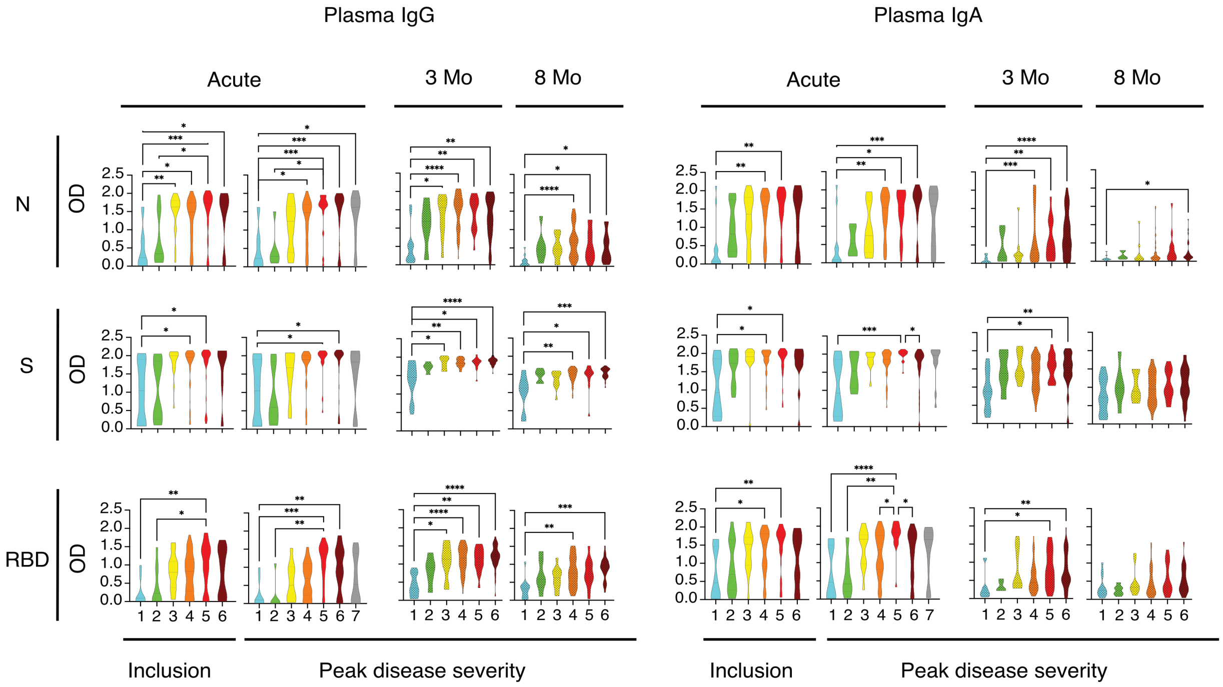


**Supplementary figure 4.** Violin plots for plasma immunoglobulins versus disease severity during the acute (n=147) and 3- (n=128) and 8-month (n=113) follow-ups. Data refer to N, S and RBD. Plots with a dotted pattern refer to follow-up data. Plots or single data points in cyan and green refer to mild disease with peak disease severity (PDS) 1 and 2, data in yellow and orange refer to moderate disease (PDS 3 and 4), data in red and cayenne refer to severe disease (PDS 5 and 6) whereas data in grey refer to patients who had a fatal outcome (PDS 7). The black lines within the plots indicate mean and interquartile range. Differences were assessed using Kruskal -Wallis with Dunn's multiple comparisons test and considered statistically significant at $p<0.05$. *$p<0.05$ ** $p<0.01$, *** $p<0.001$, **** $p<0.0001$.

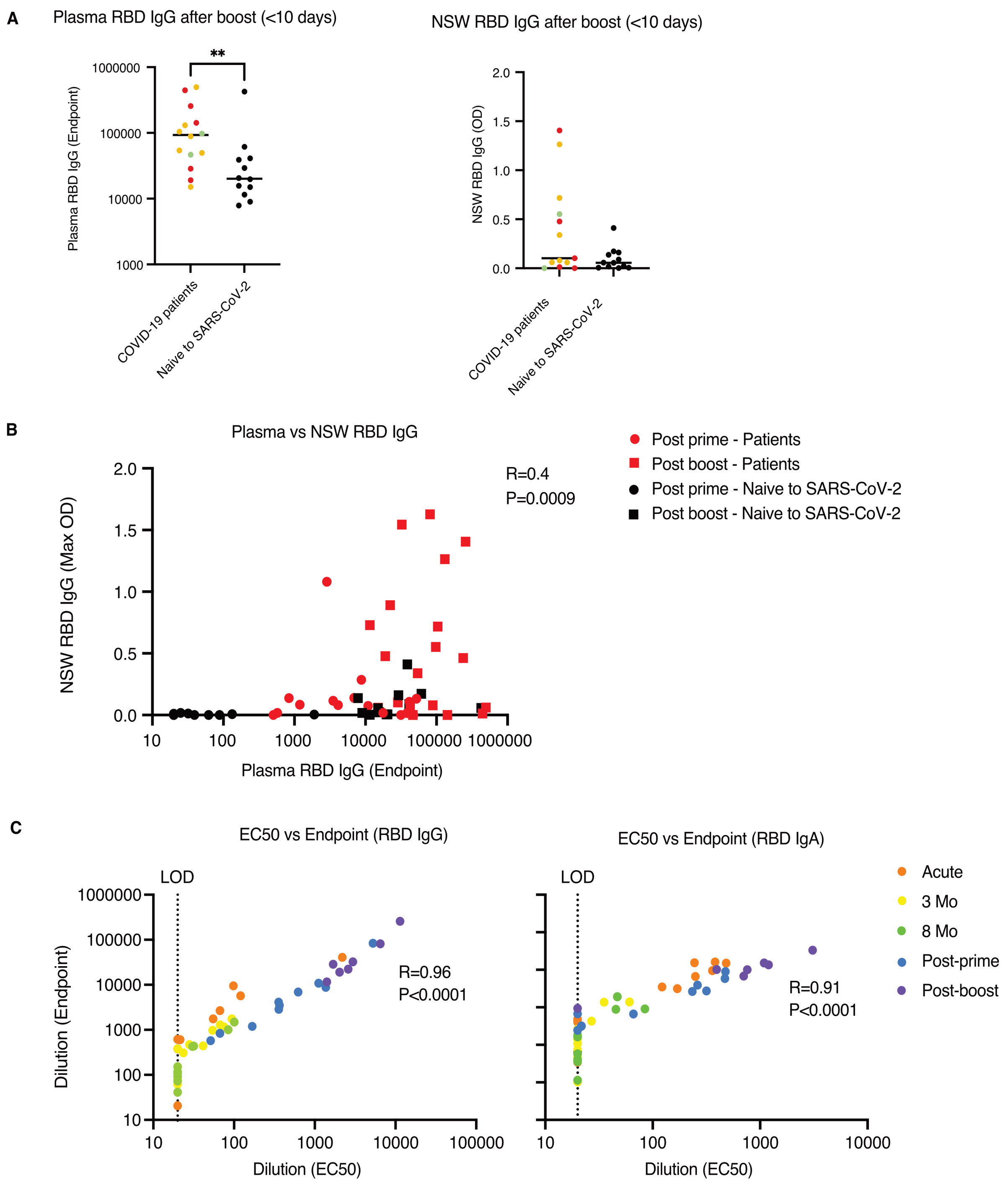


**Supplementary figure 5. (A)** Direct comparison between the levels of plasma and NSW RBD IgG between COVID-19 patients (n=14) and individuals naïve to SARS-CoV-2 (n=12) samples <10 days after boost. **(B)** Spearman correlation (n=64) between the levels of RBD IgG in plasma and in NSW after vaccination of patients (in red) and of individuals naïve to SARS-Cov-2 (in black). Circles and squares indicate the levels post-prime and post-boost respectively. **(C)** Spearman correlation between endpoint and EC50 values for the levels of plasma IgG and IgA against RBD measured in patients receiving their COVID-19 vaccination. Matched longitudinal samples (n=60) from acute, 3-month- and 8-month time points are shown together with data from after prime and after boost.

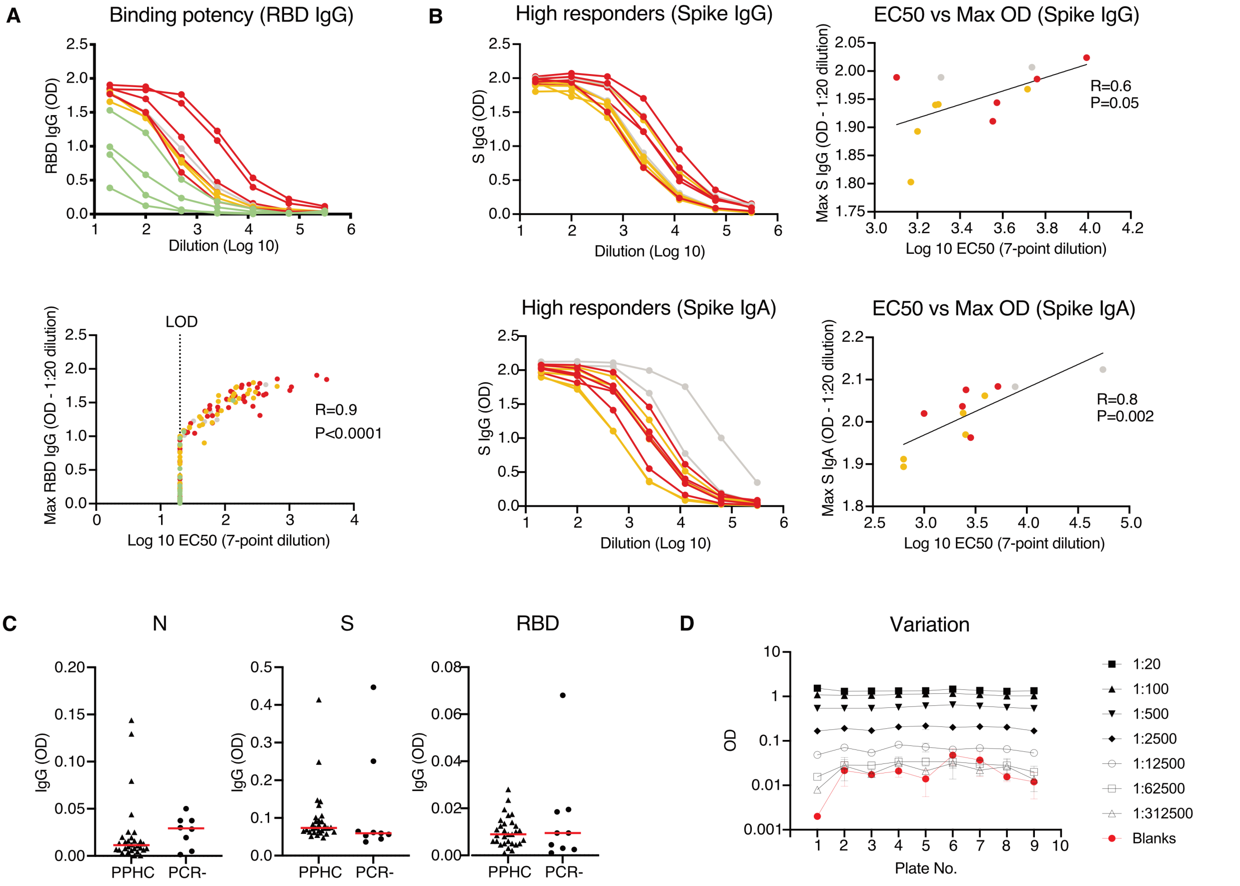


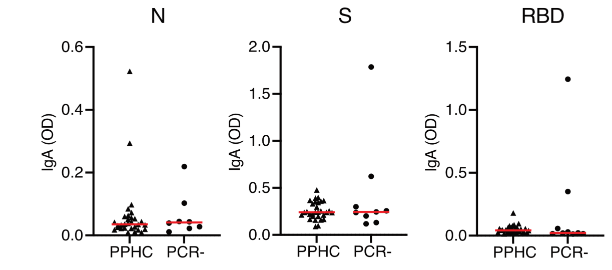


| | 1:20 | 1:100 | 1:500 | 1:2500 | 1:12500 | 1:62500 | 1:312500 | Blank |
|---|---|---|---|---|---|---|---|---|
| No of values | 9 | 9 | 9 | 9 | 9 | 9 | 9 | 9 |
| Minimum | 1.3 | 1.0 | 0.5 | 0.2 | 0.05 | 0.02 | 0.008 | 0.002 |
| 25% Perc. | 1.3 | 1.0 | 0.5 | 0.2 | 0.05 | 0.02 | 0.02 | 0.013 |
| Median | 1.3 | 1.1 | 0.6 | 0.2 | 0.06 | 0.03 | 0.02 | 0.02 |
| 75% Perc. | 1.4 | 1.1 | 0.6 | 0.2 | 0.07 | 0.03 | 0.03 | 0.03 |
| Maximum | 1.5 | 1.2 | 0.6 | 0.2 | 0.08 | 0.03 | 0.03 | 0.05 |
| Range | 0.23 | 0.2 | 0.1 | 0.05 | 0.03 | 0.02 | 0.02 | 0.05 |
| Mean | 1.4 | 1.1 | 0.6 | 0.2 | 0.06 | 0.03 | 0.02 | 0.02 |
| Std. Deviation | 0.08 | 0.05 | 0.04 | 0.02 | 0.01 | 0.006 | 0.008 | 0.01 |
| SE of Mean | 0.03 | 0.02 | 0.01 | 0.006 | 0.004 | 0.002 | 0.003 | 0.005 |
| Co. of variation | 5.6% | 4.7% | 7.0% | 10.2% | 16.8% | 23.0% | 36.2% | 65.4% |

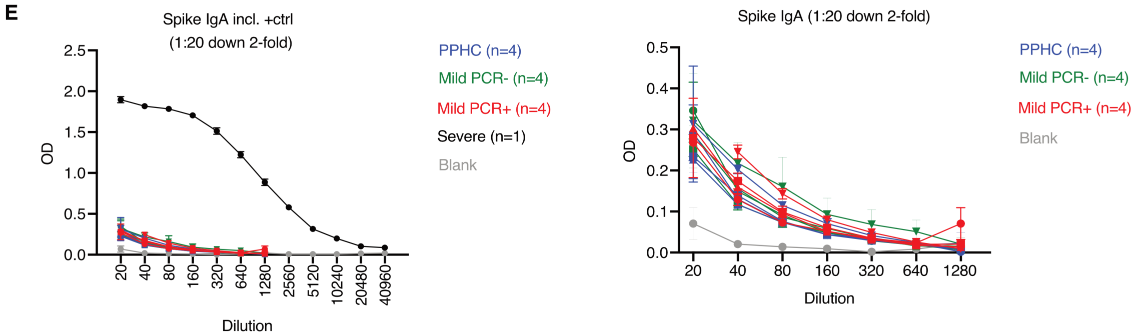


**Supplementary figure 6. (A)** Representative binding curves (n=12) for plasma IgG against RBD by ELISA in a 7-point dilution series starting from 1:20 and spearman correlation between EC50 and maximal OD at the 1:20 dilution (n=132) color-coded according to PDS. EC50 have been calculated using non-linear fit of the data to a sigmoidal curve by constraining the top of each curve to the OD of 2. LOD= limit of detection. **(B)** Representative binding curves (n=12) for plasma with high IgG and IgA against S by ELISA in a 7-point dilution series starting from 1:20 and spearman correlation between EC50 and maximal OD at the 1:20 dilution color-coded according to PDS. EC50 have been calculated as per above. **(C)** Levels of plasma IgG and IgA in pre-pandemic healthy controls (n=30) (PPHC) and people experiencing mild COVID-19 symptoms who had a negative diagnostic PCR (n=9) (PCR-). Black triangles and circles symbolize the PPHC and PCR- reference groups respectively. The red lines indicate median values. Data on N, S and RBD are shown from left to right. Mann-Whitney test was used to compare the different groups. **(D)** Calculation of the ELISA coefficient of variation with repeated measures at limiting dilutions as indicated **(E)** Binding curves on plasma IgA against S with limiting sample dilutions using selected samples with previously determined low binding titers (OD between 0.4 and 0.2). The samples were selected among PPHC (indicated in blue), PCR- (indicated in green) and mild patients (indicated in red). The two panels show the same data with the difference that a sample with high IgA titers is shown (in black) in the top panel as a reference. Blanks are indicated in grey.

**Supplementary table 1:** Regression model with splines. The numbers for each antibody type in each severity group indicate the change as compared with the mild group. Intercept represents the baseline, Spline1 refers to the first 15 days, Spline2 to the interval between 15 and 50 days and Spline3 refers to 50+ days. The splines were set based on a visual inspection done with the help of kernel smoothed graphs.

| | N IgG | S IgG | RBD IgG | N IgA | S IgA | RBD IgA |
|---|---|---|---|---|---|---|
| (Intercept) | 0.1241 | 0.5584 | 0.0074 | 0.4666 | 0.7552 | 0.5895 |
| Spline1 | 0.0351 | 0.0410 | 0.0201 | 0.0073 | 0.0615 | 0.0140 |
| Spline2 | 0.0074 | 0.0134 | 0.0097 | -0.0080 | -0.0150 | -0.0109 |
| lspline3 | -0.0035 | -0.0025 | -0.0011 | -0.0011 | -0.0015 | -0.0007 |
| Moderate | -0.4770 | -0.1007 | -0.5928 | 0.3248 | 0.4597 | -0.4080 |
| Severe | -0.5775 | -0.5508 | -0.5875 | -0.8426 | 0.3249 | -0.1874 |
| Fatal | -0.5231 | -1.3958 | -0.9058 | -0.5334 | 1.3550 | -0.0171 |
| Spline1: Moderate | 0.0858 | 0.0484 | 0.0721 | 0.0302 | -0.0171 | 0.0731 |
| Spline2: Moderate | 0.0000 | -0.0080 | 0.0026 | -0.0066 | 0.0048 | -0.0135 |
| Spline3: Moderate | -0.0017 | 0.0000 | -0.0011 | -0.0021 | -0.0013 | -0.0002 |
| Spline1: Severe | 0.0980 | 0.0776 | 0.0887 | 0.1319 | -0.0043 | 0.0731 |
| Spline2: Severe | -0.0032 | -0.0059 | -0.0009 | -0.0103 | 0.0063 | -0.0119 |
| Spline3: Severe | -0.0022 | -0.0001 | -0.0014 | -0.0028 | -0.0012 | -0.0010 |
| Spline: Fatal | 0.1020 | 0.1280 | 0.0870 | 0.0883 | -0.1003 | 0.0315 |
| Spline2: Fatal | -0.0043 | 0.0056 | 0.0248 | 0.0237 | 0.0372 | 0.0262 |
| Spline3: Fatal | NA | NA | NA | NA | NA | NA |

**Supplementary table 2:** Multivariable linear models. Estimate, standard error (SE), T and P values are shown for the two different models. In the "adjusted" model (results shown on the left), values have been adjusted for days from onset of symptoms, age, gender and comorbidities (CCI) while in the "unadjusted" model (results shown on the right), the severity group is alone used as covariate. Both models use the acute response for each group as the outcome and use the "mild" group as a reference/comparison group. Significant P values are highlighted in bold while P values that are significant in the unadjusted but not in adjusted model are highlighted in bold and Italic.

**Results outcome: Plasma N IgG acute**

| | Estimate | SE | T value | P value | Estimate | SE | T value | P value |
|---|---|---|---|---|---|---|---|---|
| Model | Adjusted | | | | Unadjusted | | | |
| (Intercept) | -0.47 | 0.34 | -1.383 | 0.169 | 0.456 | 0.143 | 3.201 | 0.002 |
| Moderate | 0.586 | 0.166 | 3.541 | **0.001** | 0.769 | 0.165 | 4.675 | **0** |
| Severe | 0.567 | 0.177 | 3.205 | **0.002** | 0.987 | 0.165 | 5.985 | **0** |
| Fatal | 0.557 | 0.23 | 2.42 | **0.017** | 0.902 | 0.229 | 3.936 | **0** |
| Days | 0.028 | 0.005 | 5.434 | 0 | NA | NA | NA | NA |
| Age | 0.013 | 0.007 | 1.923 | 0.057 | NA | NA | NA | NA |
| Gender | 0.101 | 0.116 | 0.865 | 0.389 | NA | NA | NA | NA |
| CCI | -0.089 | 0.053 | -1.688 | 0.094 | NA | NA | NA | NA |

**Results outcome: Plasma S IgG acute**

| | Estimate | SE | T value | P value | Estimate | SE | T value | P value |
|---|---|---|---|---|---|---|---|---|
| Model | Adjusted | | | | Unadjusted | | | |
| (Intercept) | 0.187 | 0.348 | 0.537 | 0.592 | 0.982 | 0.149 | 6.605 | 0 |
| Moderate | 0.476 | 0.17 | 2.804 | **0.006** | 0.65 | 0.172 | 3.786 | **0** |
| Severe | 0.301 | 0.181 | 1.657 | ***0.1*** | 0.749 | 0.172 | 4.365 | **0** |
| Fatal | 0.093 | 0.236 | 0.394 | 0.694 | 0.446 | 0.239 | 1.867 | 0.064 |
| Days | 0.032 | 0.005 | 6.018 | 0 | NA | NA | NA | NA |
| Age | 0.01 | 0.007 | 1.401 | 0.163 | NA | NA | NA | NA |
| Gender | 0.026 | 0.119 | 0.214 | 0.831 | NA | NA | NA | NA |
| CCI | -0.057 | 0.054 | -1.053 | 0.294 | NA | NA | NA | NA |

**Results outcome: Plasma RBD IgG acute**

| | Estimate | SE | T value | P value | Estimate | SE | T value | P value |
|---|---|---|---|---|---|---|---|---|
| Model | Adjusted | | | | Unadjusted | | | |
| (Intercept) | -0.412 | 0.265 | -1.554 | 0.123 | 0.238 | 0.12 | 1.99 | 0.049 |
| Moderate | 0.301 | 0.129 | 2.331 | **0.021** | 0.4 | 0.138 | 2.899 | **0.004** |
| Severe | 0.436 | 0.138 | 3.159 | **0.002** | 0.781 | 0.138 | 5.655 | **0** |
| Fatal | 0.198 | 0.18 | 1.103 | ***0.272*** | 0.447 | 0.192 | 2.325 | **0.021** |
| Days | 0.031 | 0.004 | 7.648 | 0 | NA | NA | NA | NA |
| Age | 0.007 | 0.005 | 1.197 | 0.234 | NA | NA | NA | NA |
| Gender | -0.021 | 0.091 | -0.233 | 0.816 | NA | NA | NA | NA |
| CCI | -0.043 | 0.041 | -1.032 | 0.304 | NA | NA | NA | NA |

**Results outcome: Plasma N IgA acute**

| | Estimate | SE | T value | P value | Estimate | SE | T value | P value |
|---|---|---|---|---|---|---|---|---|
| Model | Adjusted | | | | Unadjusted | | | |
| (Intercept) | -0.424 | 0.384 | -1.106 | 0.271 | 0.471 | 0.151 | 3.123 | 0.002 |
| Moderate | 0.562 | 0.187 | 3.005 | **0.003** | 0.784 | 0.174 | 4.499 | **0** |
| Severe | 0.59 | 0.2 | 2.952 | **0.004** | 0.953 | 0.175 | 5.458 | **0** |
| Fatal | 0.378 | 0.26 | 1.454 | ***0.148*** | 0.777 | 0.243 | 3.205 | **0.002** |
| Days | 0.011 | 0.006 | 1.885 | 0.062 | NA | NA | NA | NA |
| Age | 0.016 | 0.008 | 2.02 | 0.045 | NA | NA | NA | NA |
| Gender | 0.149 | 0.131 | 1.133 | 0.259 | NA | NA | NA | NA |
| CCI | -0.032 | 0.06 | -0.543 | 0.588 | NA | NA | NA | NA |

**Results outcome: Plasma S IgA acute**

| | Estimate | SE | T value | P value | Estimate | SE | T value | P value |
|---|---|---|---|---|---|---|---|---|
| Model | Adjusted | | | | Unadjusted | | | |
| (Intercept) | 0.863 | 0.285 | 3.027 | 0.003 | 1.221 | 0.11 | 11.052 | 0 |
| Moderate | 0.387 | 0.139 | 2.787 | **0.006** | 0.545 | 0.128 | 4.271 | **0** |
| Severe | 0.346 | 0.148 | 2.333 | **0.021** | 0.54 | 0.128 | 4.237 | **0** |
| Fatal | 0.32 | 0.193 | 1.655 | ***0.1*** | 0.545 | 0.178 | 3.069 | **0.003** |
| Days | 0.003 | 0.004 | 0.801 | 0.425 | NA | NA | NA | NA |
| Age | 0.01 | 0.006 | 1.643 | 0.103 | NA | NA | NA | NA |
| Gender | 0.013 | 0.098 | 0.136 | 0.892 | NA | NA | NA | NA |
| CCI | -0.05 | 0.044 | -1.119 | 0.265 | NA | NA | NA | NA |

**Results outcome: Plasma RBD IgA acute**

| | Estimate | SE | T value | P value | Estimate | SE | T value | P value |
|---|---|---|---|---|---|---|---|---|
| Model | Adjusted | | | | Unadjusted | | | |
| (Intercept) | 0.243 | 0.381 | 0.638 | 0.525 | 0.673 | 0.146 | 4.625 | 0 |
| Moderate | 0.376 | 0.186 | 2.024 | **0.045** | 0.575 | 0.168 | 3.419 | **0.001** |
| Severe | 0.489 | 0.199 | 2.461 | **0.015** | 0.709 | 0.168 | 4.216 | **0** |
| Fatal | 0.352 | 0.258 | 1.362 | ***0.175*** | 0.576 | 0.234 | 2.463 | **0.015** |
| Days | 0.003 | 0.006 | 0.584 | 0.56 | NA | NA | NA | NA |
| Age | 0.01 | 0.008 | 1.229 | 0.221 | NA | NA | NA | NA |
| Gender | 0.218 | 0.131 | 1.667 | 0.098 | NA | NA | NA | NA |
| CCI | -0.08 | 0.059 | -1.35 | 0.179 | NA | NA | NA | NA |
